# CMV seropositivity in older adults changes the T cell repertoire, but does not prevent antibody or cellular responses to SARS-CoV-2 vaccination

**DOI:** 10.1101/2022.05.27.22275673

**Authors:** Jessica A. Breznik, Angela Huynh, Ali Zhang, Lucas Bilaver, Hina Bhakta, Hannah D. Stacey, Jann C. Ang, Jonathan L. Bramson, Ishac Nazy, Matthew S. Miller, Judah Denburg, Andrew P. Costa, Dawn M. E. Bowdish, other members of the COVID-in-LTC Investigator Group

**Author notes:** **Corresponding author:** Dawn Bowdish, PhD, M. G. DeGroote Institute for Infectious Disease Research, McMaster Immunology Research Centre, McMaster University, MDCL 4020, 1280 Main Street West, Hamilton, Ontario, Canada L8S 4L8.

## Abstract

Chronic infection with human cytomegalovirus (CMV) may contribute to poor vaccine efficacy in older adults. We assessed effects of CMV serostatus on antibody quantity and quality, as well as cellular memory recall responses, after 2 and 3 SARS-CoV-2 mRNA vaccine doses, in older adults in assisted living facilities. CMV serostatus did not affect anti-Spike and anti-RBD IgG antibody levels, nor neutralization capacity against wildtype or beta variants of SARS-CoV-2 several months after vaccination. CMV seropositivity altered T cell expression of senescence-associated markers and increased T_EMRA_ cell numbers, as has been previously reported; however, this did not impact Spike-specific CD4^+^ T cell memory recall responses. CMV seropositive individuals did not have a higher incidence of COVID-19, though prior infection influenced humoral immunity. Therefore, CMV seropositivity may alter T cell composition but does not impede the durability of humoral protection or cellular memory responses after SARS-CoV-2 mRNA vaccination in older adults.

**Key Points:** CMV seropositive older adults have more EMRA and terminally differentiated T cells CMV seropositivity does not prevent antibody maintenance after SARS-CoV-2 vaccination CMV seropositivity does not impede SARS-CoV-2 vaccine T cell memory recall responses

## Introduction

Aging is associated with an increased frequency of viral respiratory infections and post-infection sequelae (1), as well as reduced efficacy and longevity of protective immunity after vaccination (2). Early in the COVID-19 pandemic, age was identified to be the most significant factor contributing to morbidity and mortality (3), and it was unclear how effective vaccines against the novel SARS-CoV-2 virus would be in older adults. In this population, vaccines that target a memory response (e.g., Herpes zoster) are often more immunogenic and effective at preventing illness than vaccines that target viral antigens that are antigenically distant from previous circulating strains (e.g., some seasonal influenza vaccines) (4-7). SARS-CoV-2 mRNA vaccines have been shown to be protective in older adults (8-10), as they are effective at generating cellular and antibody-mediated immunity (11-14). However, in older adults, immune responses after vaccination are generally quite heterogeneous, and immunity may wane faster than in younger adults (11, 13).

In older adults, both immunosenescence and inflammation are thought to influence the immunogenicity of vaccines and longevity of protective immune responses after vaccination (15, 16). Many studies have also implicated human cytomegalovirus (CMV) as a significant contributor to age-associated immune dysfunction. CMV is a common and persistent β-herpesvirus, found in ∼60-90% of adults worldwide, and seropositivity increases with age (17, 18). CMV infection is typically asymptomatic in immunocompetent individuals, but its accompanying chronic immune activation fundamentally alters immune cell composition and function. There is a consensus that CMV seropositivity has a long-term impact on the maturation and composition of immune cells, including increased numbers and prevalence of CD8^+^ T cells, with expansion of CMV-specific effector and memory cells at the expense of naïve T cells (19, 20). Age-associated immunosenescence likewise contributes to similar changes within the T cell repertoire, reducing naïve T cells and increasing memory T cell populations, which have impaired proliferation, differentiation, and effector functions (21, 22). A dysfunctional T cell repertoire may also have significant effects on B cell proliferation, differentiation, and maturation (23). Accordingly, CMV seropositivity has been implicated as an exacerbating factor in age-associated immune remodelling and inter-individual immune diversity (24-27), as well as a modifying factor that may compromise infection outcomes and quality and longevity of immune protection after vaccination (28).

Though there is little data to date, CMV reactivation in older adults has been suggested to contribute to more severe COVID-19 (29-33), as CMV seropositivity has been associated with impaired antibody production and cellular memory recall responses (34-36). However, CMV seropositivity has also been reported to enhance cellular and antibody immune responses to unrelated bacteria or viruses, through diversification of CD8^+^ T cell receptors and augmentation of basal inflammation (37-40). CMV seropositivity has, in addition, been associated with a reduced humoral response to inactivated split influenza virus vaccines (41-44) and viral-vector-based Ebola vaccines (45). Yet, a recent meta-analysis reported that there was insufficient evidence that CMV seropositive individuals have decreased antibody production after influenza vaccination (46). Recent data also show that CMV seropositivity in young adults does not affect antibody or cellular responses after vaccination with the adenovirus-based vector vaccine ChAdOx1 nCoV-19 (47). These conflicting reports may suggest context, time-, and age-dependent effects of chronic CMV infection on immune function. SARS-CoV-2 mRNA vaccines have been widely deployed in Canada, particularly in older adults. Whether CMV seropositivity impacts SARS-CoV-2 mRNA vaccine efficacy and durability of immunity is not yet known. Herein we investigated effects of CMV serostatus on vaccine-associated humoral protection and cellular memory recall responses several months after 2 and 3 doses of SARS-CoV-2 mRNA vaccines in older adults. We found that CMV serostatus does not impede antibody or cellular responses to SARS-CoV-2 vaccination in older adults.

## Methods

### Participant recruitment and blood collection

Participants in the COVID in Long-Term Care Study (covidinltc.ca) were recruited from assisted living facilities (17 nursing homes and 8 retirement homes) in Ontario, Canada, between March and December 2021. All protocols were approved by the Hamilton Integrated Research Ethics Board and other site-specific research ethics boards, and informed consent was obtained. Venous blood was drawn in anti-coagulant-free vacutainers for isolation of serum, as per standard protocols (48). Venous blood was drawn in heparin-coated vacutainers for immunophenotyping and T cell activation assays. Blood was collected at least 7 days after 2 and/or 3 mRNA vaccine doses. Participants received 2 doses of Moderna Spikevax 100 μg (mRNA-1273) or Pfizer Cominarty 30 μg (BNT162b2) as per recommended schedules, and a third mRNA vaccine dose in Fall 2021 at least 6 months from the last dose, as per Province of Ontario guidelines (49). For this manuscript, humoral and cellular data were retrospectively assessed in context of CMV serostatus from a cohort of 186 participants, 65 years of age and older. Blood was drawn after 2 and 3 mRNA vaccine doses from 47 cohort participants. Participant cohort demographics are summarized in Table 1.

**Table 1.** Participant Demographics.

|  | **CMV Seronegative**<br>**(n=74)** | **CMV Seropositive**<br>**(n=159)** | **Statistical**<br>**Assessment** |
| --- | --- | --- | --- |
| Age<br>(median ± SD; range) | 85 ± 6.8 years<br>(65-93) | 86 ± 7.7 years<br>(65-101) | p=0.1265* |
| Sex (frequency) | 67.6% female (n=50) | 69.8% female (n=111) | p=0.7618** |
| Sample Size<br>Post-Dose 2 | n=51 | n=108 | .-- |
| 2<sup>nd</sup> Dose Vaccine<br>Combination<br>(frequency) | 47.1% Moderna-Moderna<br>(n=24)<br>52.9% Pfizer-Pfizer (n=27) | 35.8% Moderna-Moderna (n=39)<br>60.6% Pfizer-Pfizer (n=66)<br>2.75% Pfizer-Moderna (n=3) | p=0.2968** |
| Days Between Dose 1<br>and Dose 2<br>(mean ± SD) | 32.2 ± 20.8 days | 30.4 ± 17.9 days | p=0.6697* |
| Days Since 2<sup>nd</sup> Dose to<br>Blood Collection<br>(mean ± SD) | 179.8 ± 50.1 days | 175.4 ± 64.1 days | p=0.9041* |
| Sample Size<br>Post-Dose 3*** | n=23 (17 repeated) | n=51 (30 repeated) | .-- |
| 3<sup>rd</sup> Dose Vaccine<br>Combination<br>(frequency) | 65.2% Moderna-Moderna-Moderna (n=15)<br>34.8% Pfizer-Pfizer-Pfizer (n=8) | 56.9% Moderna-Moderna-Moderna<br>(n=29)<br>43.1% Pfizer-Pfizer-Pfizer (n=22) | p=0.6115** |
| Days Between Dose 2<br>and Dose 3<br>(mean ± SD) | 203.9 ± 21.0 days | 211.7 ± 12.1 days | p=0.2689* |
| Days Since 3<sup>rd</sup> Dose to<br>Blood Collection<br>(mean ± SD) | 82.0 ± 17.3 days | 83.7 ± 11.5 days | p=0.7597* |
| Prior COVID-19**** | 37.8% (n=28) | 34.4% (n=55) | p=0.6609* |
| Only positive for<br>nasopharyngeal PCR<br>test | 28.6% (n=8) | 36.4% (n=20) | p>0.6244** |
| Only positive for anti-NC<br>antibodies | 39.3% (n=11) | 40.0% (n=22) | p>0.9999** |
| Positive for PCR test<br>and anti-NC antibodies | 32.1% (n=9) | 23.6% (n=13) | p=0.7977** |

\*Student's t test with Welch's correction or Mann-Whitney U-test by normality.
\*\*Fisher's exact test. Comparisons for vaccines post-dose 2 were calculated for Moderna-Moderna vs Pfizer-Pfizer.
\*\*\* 47 participants had blood drawn post-dose 2 and post-dose 3.
\*\*\*\* Participants were considered 'positive' for prior COVID-19 infection if they had a recorded positive nasopharyngeal PCR test at a date prior to blood collection and/or seropositivity for anti-nucleocapsid (NC) IgG or IgA antibodies at time of blood collection for antibody and cellular assays.

### Determination of CMV serostatus

CMV seropositivity was determined by enzyme-linked immunosorbent assay (ELISA) with the Human Anti-Cytomegalovirus IgG ELISA Kit (CMV) (#ab108639, Abcam) as per manufacturer’s instructions. Serum samples from a participant’s first blood draw were diluted 1:40 and assessed in duplicate. Samples with a CMV IgG index above or equal to the positive standard were classified as CMV seropositive, whereas samples with a CMV IgG index less than the positive standard were classified as CMV seronegative.

### Whole blood immunophenotyping

Circulating immune cell populations were quantitated in whole blood using fluorophore-conjugated monoclonal antibodies by multicolour flow cytometry with a CytoFLEX LX (4 laser, Beckman Coulter), as per standard protocols (48, 50, 51). Count-Bright™ absolute counting beads (#C36950, Invitrogen Life Technologies) were used to determine absolute cell counts. Data was analyzed with FlowJo V10.8.1 (TreeStar, Inc.), following a previously published gating strategy to identify T cell populations (50). Five main subsets of human CD8^+^ and CD4^+^ T cells (Naïve (N), Central Memory (CM), Effector Memory (EM), Effector Memory re-expressing CD45RA (EMRA), and terminally differentiated (TD)) were identified by their expression of CD45RA, CCR7, CD28 and/or CD57. CD8_N_ and CD4_N_ were classified as CD45RA^+^CCR7^+^; CD8_CM_ and CD4_CM_ as CD45RA^-^CCR7^+^; CD8_EM_ and CD4_EM_ as CD45RA^-^ CCR7^-^, CD8_EMRA_ and CD4_EMRA_ as CD45RA^+^CCR7^-^; and CD8_TD_ and CD4_TD_ as CD45RA^+^CCR7^-^ CD28^-^CD57^+^, as per standard protocols (52).

### Assessment of T cell memory responses to SARS-CoV-2 Spike by Activation Induced Marker (AIM) assay

Antigen-specific T cell recall responses were evaluated by AIM assay as per established protocols (50). Each participant sample was stimulated with a Spike glycoprotein SARS-CoV-2 peptide pool (1 μg/mL) containing overlapping peptides of the complete immunodominant sequence domain (#130-126-701; Miltenyi Biotec), and influenza hemagglutinin (HA) peptides (4 μL of 0.12 mcg/μL HA; AgriFlu, Alfuria® Tetra Inactivated Influenza Vaccine 2020-2021 season, Seqirus, UK). A negative media control (unstimulated) and positive stimulation control (polyclonal stimulation with CytoStim™ (0.5 μL/well, #130-092-173; Miltenyi Biotec)) were included with each sample. For each of these four conditions, 100 μL of heparinized venous blood was incubated with an equal volume of Iscove’s Modified Dulbecco’s Medium with GlutaMAX™ Supplement (#31980030, Invitrogen Life Technologies) for 44 h in 96-well flat bottom plates at 37°C. Samples were stained with fluorophore-conjugated monoclonal antibodies and assessed with a CytoFLEX LX (4 laser, Beckman Coulter, Brea, CA, USA) as previously described (50). Data was analyzed with FlowJo, following a previously published gating strategy to identify AIM^+^ T cells (50). Activated T cells (AIM-positive) were identified by their co-expression of CD25 and CD134 (OX40) on CD4^+^ T cells (53, 54), and co-expression of CD69 and CD137 (4-1BB) on CD8^+^ T cells (55). Samples with a T cell count of at least 20 events and ≥2-fold above the unstimulated sample (negative control; i.e., stimulation index ≥2), were defined as AIM-positive. Expression of CXCR3 (CD183; Brilliant Violet 421, #353716, BioLegend), CCR4 (CD194; Brilliant Violet 605, #359418, BioLegend,) and/or CCR6 (CD196; Brilliant Violet 785, #353422, BioLegend) was used to identify Th1 (CXCR3^+^CCR6^-^CCR4^-^), Th2 (CXCR3^-^CCR4^+^CCR6^-^), and Th17 (CXCR3^-^CCR4^+^CCR6^+^) AIM^+^CD4^+^ T cell subsets.

### Measurements of anti-SARS-CoV-2 antibodies and neutralizing capacity

Serum anti-SARS-CoV-2 Spike protein and receptor binding domain (RBD) IgG, IgA and IgM antibodies were measured by a validated ELISA as previously described (50, 56), with assay cut-off 3 standard deviations above the mean of a pre-COVID-19 population from the same geographic region. Antibody neutralization capacity was assessed by cell culture assays with Vero E6 (ATCC CRL-1586) cells and live SARS-CoV-2, with data reported as geometric microneutralization titers at 50% (MNT_50_), which ranged from below detection (MNT_50_ = 5; 1:10 dilution) to MNT_50_ = 1280 (56). Antibody neutralization was measured against the ancestral strain of SARS-CoV-2 and the beta variant of concern (B.1.351). The beta variant was obtained through BEI Resources, NIAID, NIH: SARS-Related Coronavirus 2, Isolate hCoV-19/South Africa/KRISP-K005325/2020, NR-54009, contributed by Alex Sigal and Tulio de Oliveira.

### Determination of prior SARS-CoV-2 infection

Due to demand for and limits on availability of testing, participants were not consistently tested for COVID-19 by nasopharyngeal swab PCR assay, even if they were symptomatic. Serological testing for anti-nucleocapsid SARS-CoV-2 IgG and IgA antibodies was performed on all collected samples by the ELISA described above (50, 56), using assay wells coated with 2 μg/ml of nucleocapsid antigen (Jackson ImmunoResearch Laboratories, Inc). Participants were identified to have had COVID-19 if they were seropositive for IgG or IgA anti-nucleocapsid antibodies, and/or had a documented positive nasopharyngeal PCR test prior to any blood collection, as summarized in Table 1.

### Statistical analysis

Statistical analyses were conducted using GraphPad Prism version 9 (San Diego, CA, USA). Two-group comparisons of dose and CMV seropositivity or prior COVID-19 and CMV seropositivity were assessed by two-way ANOVA. Differences between CMV seropositive and seronegative group antibody levels, antibody neutralization capacity, T cell immunophenotype, and T cell memory recall responses, were assessed by Student’s t-test with Welch’s correction or Mann-Whitney U-test, according to data normality. P values are reported as two-tailed and p values less than 0.05 were considered significant.

## Results

### Participant demographics

Serum anti-CMV IgG antibodies were measured by ELISA, and 69.4% (n = 129/186) of participants were found to be CMV seropositive. Age and sex distribution were similar between seropositive (median 85±6.8 years, 67.6% female) and seronegative (median 86±7.7 years, 69.8% female) participants (Table 1). Blood samples were collected at a median of 179.8 days (CMV seronegative) and 175.4 days (CMV seronegative) after 2 doses of Moderna Spikevax 100 μg (mRNA-1273) or Pfizer Cominarty 30 μg (BNT162b2) administered as per manufacturer-recommended schedules. In Ontario, Canada, 3^rd^ dose vaccinations were recommended for older adults in congregate living beginning in August 2021, if they were more than 6 months post-2^nd^ vaccinations (57). Participants received 3^rd^ doses in August-September 2021, and blood samples were collected at a median of 82.0 days (CMV seronegative) or 83.7 days (CMV seropositive) after 3^rd^ doses. Participants were classified as having had a previous SARS-CoV-2 infection if they had a documented positive PCR test and/or were seropositive for IgG or IgA nucleocapsid antibodies. A positive nasopharyngeal PCR test and/or serum anti-nucleocapsid IgG or IgA antibodies were reported in 37.8% of CMV seronegative participants and 34.4% of CMV seropositive participants.

### CMV seropositivity does not impede anti-SARS-CoV-2 antibody production or neutralization in older adults

To assess effects of CMV serostatus on antibody responses after SARS-CoV-2 vaccination, serum anti-SARS-CoV-2 Spike and RBD IgG, IgA, and IgM antibody levels were measured by ELISA (Figure 1; Table 2). The number of responders (i.e., individuals with antibodies above the threshold limit of detection) significantly increased between post-2^nd^ and post-3^rd^ dose measurements of anti-Spike IgG, IgA, and IgM antibodies, as well as anti-RBD IgG antibodies, but not anti-RBD IgA or IgM antibodies (Table 2). For example, 5-7 months after second dose vaccinations, anti-Spike IgG antibodies were detected in 88.7% of participants, and anti-RBD IgG antibodies were detected in 63.5% of participants. Approximately 3 months after the third vaccine dose, 97.3% and 94.6% of participants had detectable anti-Spike IgG and anti-RBD IgG, respectively. CMV seropositivity did not impact the frequency of responders for anti-Spike and anti-RBD IgG, IgA, and IgM antibodies. Accordingly, two-group analyses showed a main effect of vaccine dose, but not CMV serostatus, on serum anti-Spike IgG (Figure 1A) and anti-RBD IgG (Figure 1D) antibodies, anti-Spike IgA (Figure 1B) and anti-RBD IgA (Figure 1E) antibodies, as well as anti-Spike IgM (Figure 1C) antibodies. There were no significant main effects of vaccine dose or CMV serostatus on anti-RBD IgM (Figure 1F) antibodies. Therefore, CMV seropositivity does not impede maintenance of antiviral antibodies several months after two or three 3 doses of SARS-CoV-2 mRNA vaccines.

**Table 2.** Responders to vaccination by CMV serostatus – antibodies.

| Antibody | Vaccine Doses | Total Responder Frequency |  |  | Responder Frequency by CMV Seropositivity |  |  |  |  |
| --- | --- | --- | --- | --- | --- | --- | --- | --- | --- |
|  |  | n | Frequency (%) | *p*<br>(Fisher's exact test) | Seronegative |  | Seropositive |  | *p*<br>(Fisher's exact test) |
|  |  |  |  |  | n | Frequency (%) | n | Frequency (%) |  |
| anti-Spike IgG | 2 | 141/159 | 88.7 | 0.0415 | 47/51 | 92.2 | 94/108 | 87.0 | 0.4287 |
|  | 3 | 72/74 | 97.3 |  | 21/23 | 91.3 | 51/51 | 100 | 0.0937 |
| anti-Spike IgA | 2 | 98/159 | 61.6 | <0.0001 | 33/51 | 64.7 | 65/108 | 60.2 | 0.6053 |
|  | 3 | 71/74 | 95.9 |  | 21/23 | 91.3 | 50/51 | 98.0 | 0.2264 |
| anti-Spike IgM | 2 | 20/159 | 12.6 | <0.0001 | 8/51 | 15.7 | 12/108 | 11.1 | 0.4476 |
|  | 3 | 28/74 | 37.8 |  | 8/23 | 34.8 | 20/51 | 39.2 | 0.7992 |
| anti-RBD IgG | 2 | 101/159 | 63.5 | <0.0001 | 34/51 | 66.7 | 67/108 | 62.0 | 0.6011 |
|  | 3 | 70/74 | 94.6 |  | 21/23 | 91.3 | 49/51 | 96.1 | 0.5837 |
| anti-RBD IgA | 2 | 35/159 | 22.0 | 0.0760 | 12/51 | 23.5 | 23/108 | 21.3 | 0.8380 |
|  | 3 | 25/74 | 33.8 |  | 8/23 | 13.0 | 17/51 | 33.3 | >0.9999 |
| anti-RBD IgM | 2 | 6/159 | 3.77 | >0.9999 | 3/51 | 5.88 | 3/108 | 2.78 | 0.3863 |
|  | 3 | 3/74 | 4.05 |  | 1/23 | 4.35 | 2/51 | 3.92 | >0.9999 |

**Figure 1.**
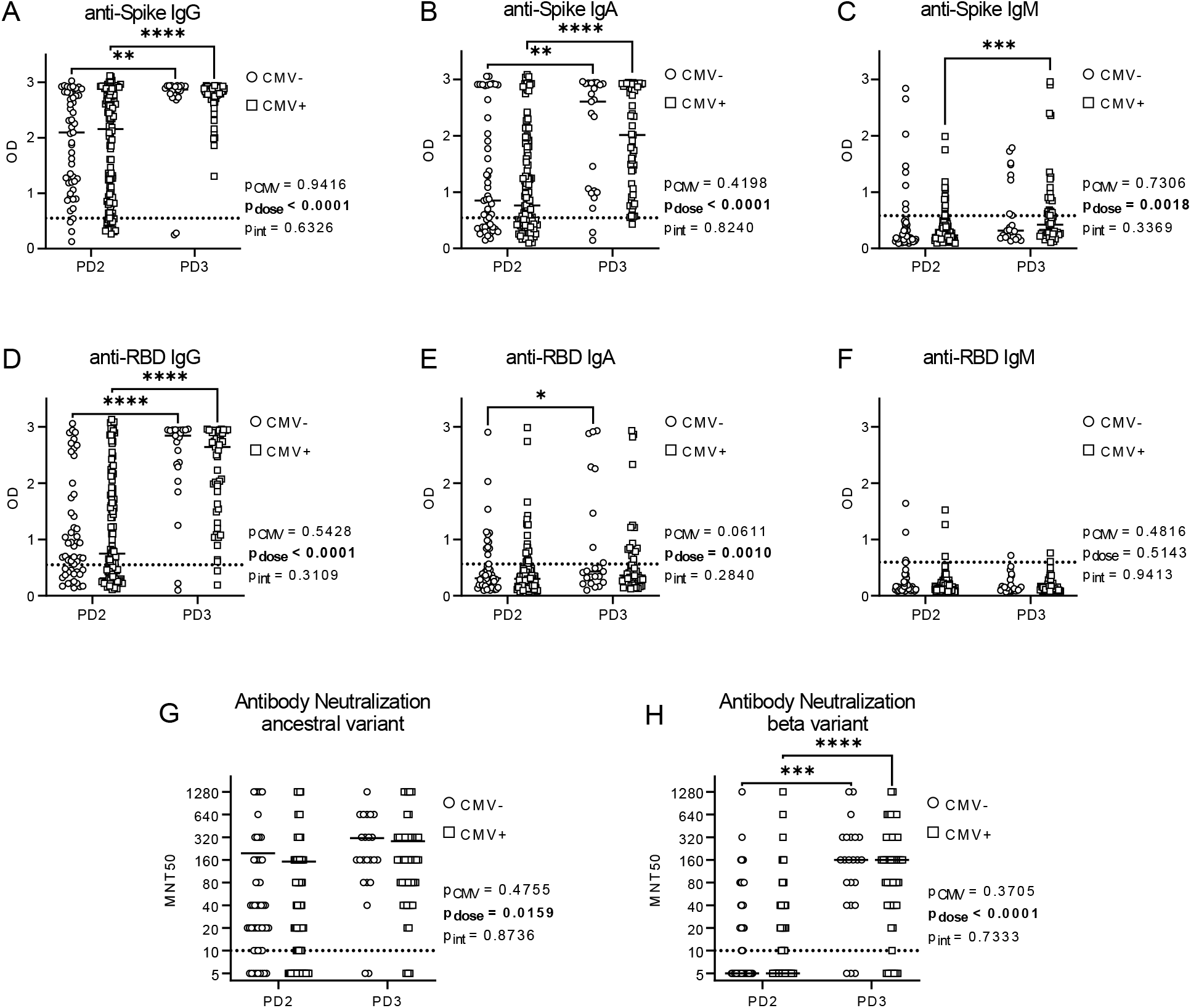
Antibodies and neutralization capacity by CMV serostatus after two and three COVID-19 vaccines in older adults. Serum SARS-CoV-2 anti-Spike and anti-RBD antibodies were detected by ELISA, and antibody neutralization capacity was assessed by MNT50 with live SARS-CoV-2 virus, post-dose 2 (PD2) and post-dose 3 (PD3) vaccination in CMV seronegative (-ve) and CMV seropositive (+ve) individuals. Anti-Spike antibodies: IgG (A), IgA (B) and IgM (C). Anti-RBD antibodies: IgG (D), IgA (E), and IgM (F). Antibody neutralization capacity was assessed against ancestral (G) and beta variant (H) SARS-CoV-2. CMV-PD2 n=51; CMV-PD3 n=23; CMV+PD2 n=108; CMV+PD3 n=51. Dotted lines indicate the threshold of detection. Each data point indicates an individual participant, with the center line at the median. Associations between CMV serostatus and vaccine dose were assessed by two-way ANOVA, with Tukey’s test post-hoc analysis of significant main effects. \**p<0*.*05*, \*\**p<0*.*01, ***p<0*.*001, ****p<0*.*0001*.

To examine potential effects of CMV serostatus on antibody function, serum antibody neutralization capacity was assessed by MNT50 assays against live ancestral (wildtype) and beta variant SARS-CoV-2 (Figure 1G-H). Vaccines were designed against the wildtype virus, whereas the beta variant contains mutations that confer increased transmissibility and immune evasion (58). Neutralization of ancestral and beta variant SARS-CoV-2 ranged from below the detection limit to MNT50=1280, though mean neutralization was consistently higher against the ancestral virus compared to the beta variant after 2 and 3 vaccine doses. Neutralization capacity was similar between CMV seropositive and seronegative individuals against both ancestral and beta variant SARS-CoV-2, though there was a main effect of dose on neutralization capacity. In particular, significant increases in serum antibody neutralization were observed against the beta variant between 2^nd^ and 3^rd^ dose vaccinations. Anti-Spike and anti-RBD IgG levels moderately correlated with antibody neutralization capacity, irrespective of CMV serostatus (Table 3). As well, modest correlations were generally observed between ancestral SARS-CoV-2 neutralization capacity and anti-Spike and anti-RBD IgA antibodies in both CMV seropositive and CMV seronegative participants. Therefore, CMV seropositivity does not compromise maintenance of vaccine-elicited antibody neutralization of SARS-CoV-2 several months after two or three vaccine doses.

**Table 3.** Associations of antibody and neutralization responses after vaccination by CMV serostatus.

|  | Antibody | MNT50 wildtype |  | MNT50 beta |  |
| --- | --- | --- | --- | --- | --- |
|  |  | CMV-ve | CMV+ve | CMV-ve | CMV+ve |
| Post-Dose 2 | IgG Spike | ****<br>$r = 0.8367$ | ****<br>$r = 0.8316$ | ****<br>$r = 0.6963$ | ****<br>$r = 0.7541$ |
| | IgG RBD | ****<br>$r = 0.7394$ | ****<br>$r = 0.8291$ | ****<br>$r = 0.6848$ | ****<br>$r = 0.6981$ |
| | IgA Spike | ****<br>$r = 0.6222$ | ****<br>$r = 0.6152$ | ****<br>$r = 0.6402$ | ****<br>$r = 0.5254$ |
| | IgA RBD | **<br>$r = 0.4311$ | ****<br>$r = 0.4572$ | **<br>$r = 0.4813$ | ***<br>$r = 0.3639$ |
| Post-Dose 3 | IgG Spike | ***<br>$r = 0.6923$ | ***<br>$r = 0.5004$ | ***<br>$r = 0.6545$ | ***<br>$r = 0.5038$ |
| | IgG RBD | ****<br>$r = 0.8207$ | ****<br>$r = 0.5770$ | ****<br>$r = 0.7475$ | ****<br>$r = 0.6926$ |
| | IgA Spike | **<br>$r = 0.6324$ | *<br>$r = 0.3228$ | *<br>$r = 0.4550$ | ns |
| | IgA RBD | ****<br>$r = 0.7750$ | *<br>$r = 0.3081$ | ***<br>$r = 0.7051$ | *<br>$r = 0.3575$ |

Spearman's correlation coefficient is reported.

As serum was collected from some participants after both 2^nd^ and 3^rd^ vaccine doses, these paired data were also assessed independently (Supplementary Figure 1). Consistent with our pooled participant data, there were main effects of vaccine dose but not CMV serostatus on anti-Spike and anti-RBD IgG and IgA serum antibody levels. We did observe an interaction between CMV serostatus and vaccine dose on anti-RBD IgM measurements by intra-individual analysis, but most participants had antibody levels below the threshold. Intra-individual analyses also showed that the number of vaccine doses, but not CMV serostatus, had a significant effect on antibody neutralization capacity against wildtype and beta variant SARS-CoV-2. Therefore, CMV seropositivity does not significantly impact intra-individual changes in antibody levels or neutralization capacity between 2 and 3 doses of SARS-CoV-2 mRNA vaccines.

We next considered post-dose 2 and post-dose 3 vaccination antibody measurements in context of prior SARS-CoV-2 infection (Figure 2). As summarized in Table 1, incidence of COVID-19 (prior to blood collections) was similar between CMV seronegative and CMV seropositive participants. We observed a main effect of prior SARS-CoV-2 infection on anti-Spike and anti-RBD IgG, IgA, and IgM antibodies after 2 doses of mRNA vaccines, as well as anti-Spike and anti-RBD IgG and IgA, but not IgM, serum antibodies after 3 vaccine doses. We observed an interaction between prior COVID-19 and CMV serostatus for anti-Spike and anti-RBD IgA antibodies post-dose 2 and post-dose 3, and IgM antibodies post-dose 2. There was a main effect of CMV serostatus on anti-Spike IgM antibodies, but most individuals had levels below the detection threshold. We in addition observed main effects of prior SARS-CoV-2 infection, but not CMV serostatus, on antibody neutralization of ancestral SARS-CoV-2 after 2 and 3 vaccine doses, and the beta variant after 2, but not 3, vaccine doses. Collectively, these data indicate that CMV serostatus does not appear to have a major impact on the longevity of circulating anti-Spike or anti-RBD IgG antibodies, or total serum antibody neutralization capacity, after SARS-CoV-2 infection or vaccination.

**Figure 2.**
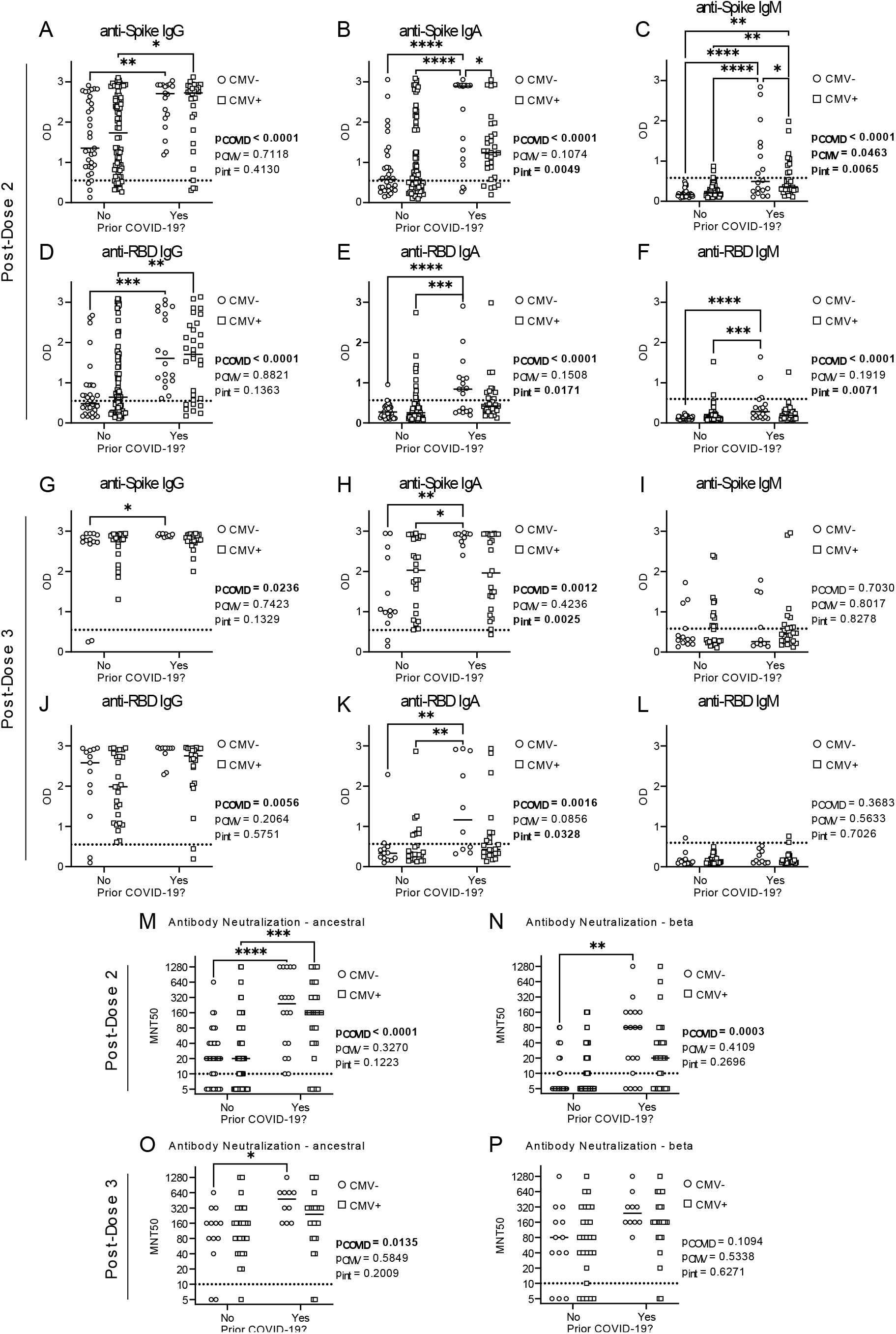
Antibodies and neutralization capacity by CMV serostatus and prior SARS-CoV-2 infection after two and three COVID-19 vaccines in older adults. SARS-CoV-2 anti-Spike and anti-RBD antibodies were measured in serum of CMV seronegative (CMV-) and CMV seropositive (CMV+) individuals by ELISA, and serum antibody neutralization capacity was assessed by MNT50 with live SARS-CoV-2 virus. Data are stratified by prior SARS-CoV-2 infection history. Post-dose 2: anti-Spike IgG (A), IgA (B) and IgM (C) antibodies, and anti-RBD IgG (D), IgA (E), and IgM (F) antibodies. Post-dose 3: anti-Spike IgG (G), IgA (H) and IgM (I) antibodies, and anti-RBD IgG (J), IgA (K), and IgM (L) antibodies. Post-dose 2 SARS-CoV-2 neutralization: ancestral (M) and beta variant (N). Post-dose 3 SARS-CoV-2 neutralization: ancestral (O) and beta variant (P). PD2: CMV-No n=33; CMV-Yes n=18; CMV+No n=77; CMV+Yes n=31. PD3: CMV-No n=13; CMV-Yes n=10; CMV+No n=27; CMV+Yes n=24. Dotted lines indicate the threshold of detection. Each data point indicates an individual participant, with the center line at the median. Associations between CMV serostatus and prior COVID-19 were assessed by two-way ANOVA, with Tukey’s test post-hoc analysis of significant main effects and interactions. *\*p<0*.*05, **p<0*.*01, ***p<0*.*001, ****p<0*.*0001*.

### CMV serostatus influences peripheral CD4^+^ and CD8^+^ T cell immunophenotype in older adults

To examine the impact of CMV seropositivity on the T cell repertoire, whole blood CD4^+^ and CD8^+^ T cell composition was quantitated, and their surface expression of CD28 and CD57 was measured, by flow cytometry (Figure 3; Supplementary Figure 2). Chronic T cell activation and an altered T cell repertoire are characteristics of CMV seropositivity (20, 38, 59). Accordingly, CMV seropositive individuals had significant changes to their peripheral blood T cell composition. We found no changes in numbers of circulating total leukocytes, total CD4^+^ T cells, or CD4_N_, CD4_EM_, CD8_N_, or CD8_EM_ T cell populations by CMV serostatus. However, CMV seropositivity increased numbers of total CD8^+^ T cells, as well as CD4_EMRA_, CD4_TD_, CD8_EMRA_, and CD8_TD_ T cells, and decreased numbers of CD4_CM_ and CD8_CM_ T cells.

**Figure 3.**
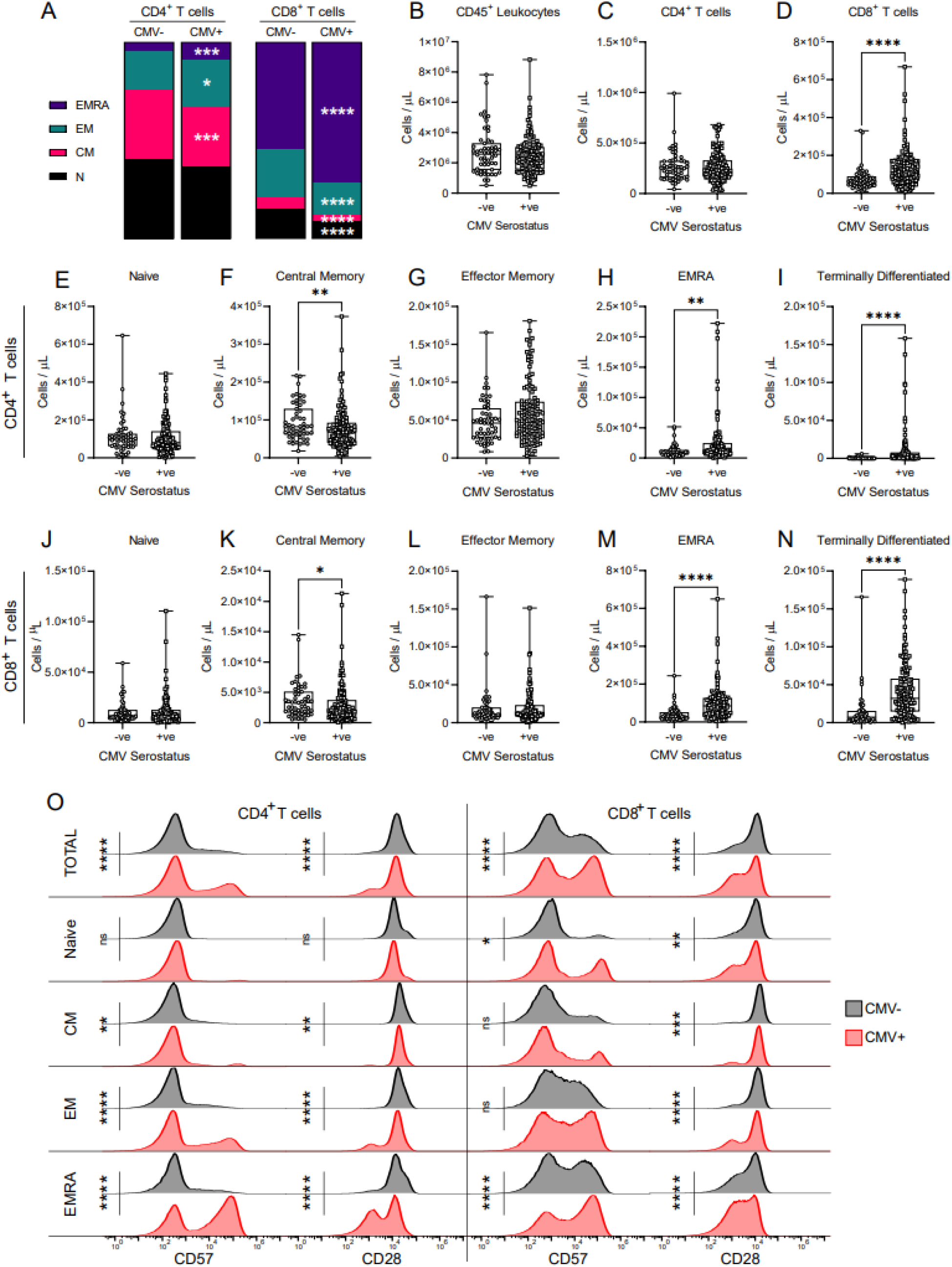
Effect of CMV serostatus on the circulating T cell repertoire in older adults. T cell populations in whole blood were assessed by flow cytometry in CMV seronegative (-ve) and CMV seropositive (+ve) individuals. (A) Relative prevalence of CD4^+^ and CD8^+^ T cell subsets by CMV serostatus (also see Supplementary Figure 2). Absolute cell counts of: (B) total leukocytes; (C) total CD4^+^ T cells; (D) CD8^+^ T cells. Absolute cell counts of CD4^+^ T cells: (E) naïve; (F) central memory; (G) effector memory; (H) EMRA; (I) terminally differentiated. Absolute cell counts of CD8^+^ T cells: (J) naive; (K) central memory; (L) effector memory; (M) EMRA; (N) terminally differentiated. (O) Surface geometric mean expression of CD57 and CD28 on CD4^+^ and CD8^+^ T cells by CMV serostatus. Blood was assessed either post-dose 2 or post-dose 3 for each participant. CMV-ve n=56; CMV+ve n=128. Each data point in B-N indicates an individual participant, and data are presented as box and whisker plots, minimum to maximum, with the center line at the median. The surface marker expression in O was visualized by concatenating uncompensated events in FlowJo for each participant and indicated T cell population grouped according to CMV serostatus, and then geometric mean fluorescence intensity expression data of each CMV group was overlaid onto the same histogram plot. Associations between T cell subsets and CMV serostatus were assessed by Student’s t test with Welch’s correction or Mann-Whitney U test, according to normality. *\*p<0*.*05, **p<0*.*01, ***p<0*.*001, ****p<0*.*0001*.

CD28 is a co-stimulatory molecule that contributes to TCR-antigen-mediated activation of T cells, while CD57 is a marker of terminally differentiated T cells as well as an indicator of immune senescence (60). Repeated T cell activation is associated with upregulation of CD57 and a reduction in CD28 expression (61-63). Consistent with these prior data, comparisons of CD28 and CD57 expression on T cell populations by CMV serostatus in our cohort of older adults (Figure 3O) revealed increased CD57 expression and reduced CD28 expression on total CD4^+^ and CD8^+^ T cell populations, as well as more specifically CD4_CM_, CD4_EM_, CD4_EMRA_, CD8_N_, and CD8_EMRA_ T cells, in CMV seropositive individuals. Expression of CD28 was also decreased on CD8_CM_ and CD8_EM_ cells of CMV seropositive individuals, though their expression of CD57 was not influenced by CMV serostatus. CMV serostatus did not alter CD57 or CD28 expression on CD4_N_ T cells.

As even mild COVD-19 can have lasting effects on immune cell composition (50), we also considered combined effects of prior SARS-CoV-2 infection and CMV serostatus on T cell composition (Figure 4; Supplementary Figure 2). Prior SARS-CoV-2 infection was associated with lower total leukocyte counts, and an interaction was observed between CMV serostatus and prior COVID-19 that influenced numbers of CD8_TD_ cells. But there were otherwise no significant main effects of prior SARS-CoV-2 infection on absolute cell numbers nor the prevalence of the assessed CD4^+^ or CD8^+^ T cell populations.

**Figure 4.**
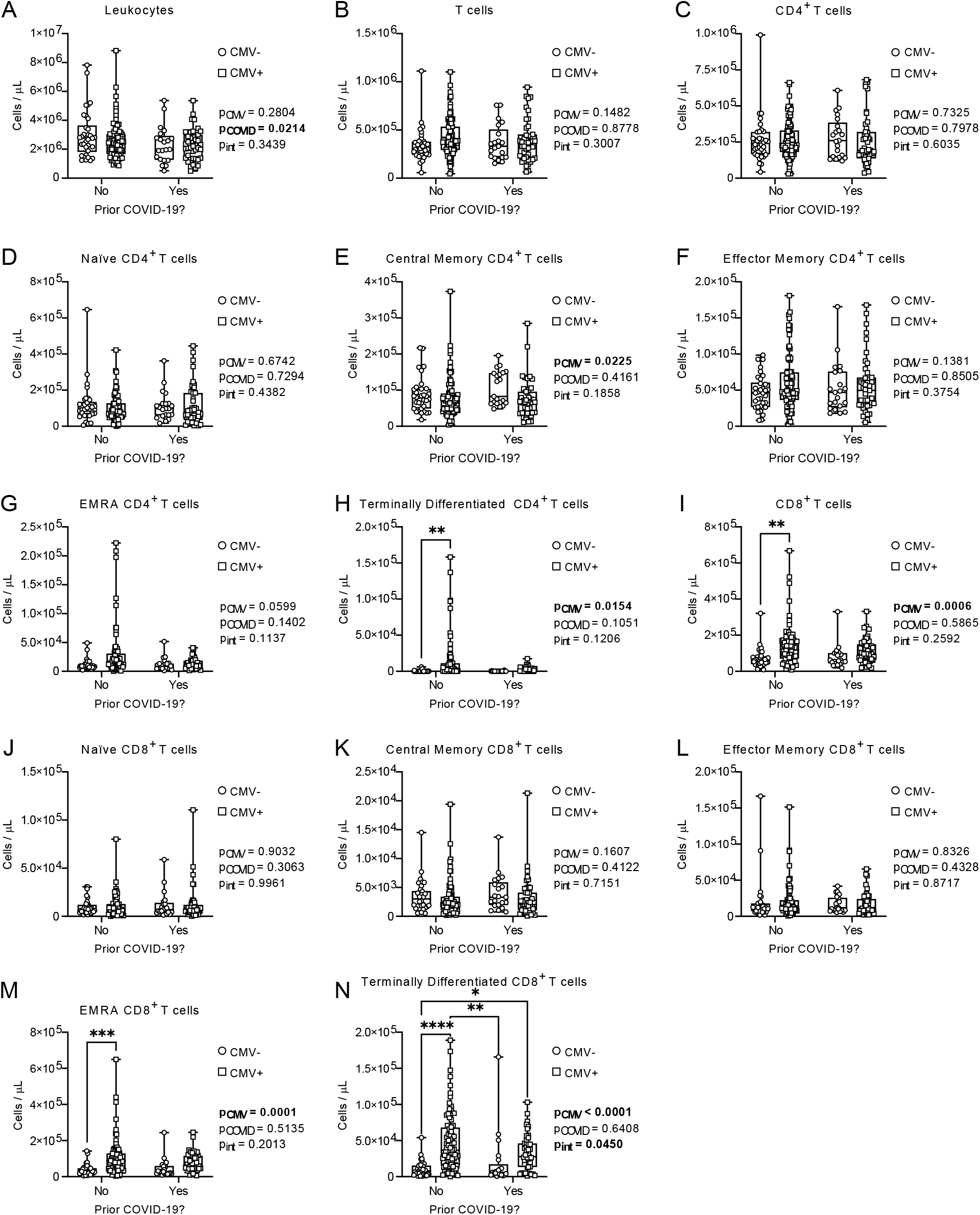
Effects of CMV serostatus and prior COVID-19 on the circulating T cell repertoire in older adults. T cell populations in whole blood were assessed by flow cytometry in CMV seronegative (CMV-) and CMV seropositive (CMV+) individuals. Data are stratified by prior SARS-CoV-2 infection history. Absolute cell counts of: (A) total leukocytes; (B) total T cells. Absolute cell counts of CD4^+^ T cells: (C) total; (D) naïve; (E) central memory; (F) effector memory; (G) EMRA; (H) terminally differentiated. Absolute cell counts of CD8^+^ T cells: (I) total; (J) naive; (K) central memory; (L) effector memory; (M) EMRA; (N) terminally differentiated. Blood was assessed either post-dose 2 or post-dose 3 for each participant. CMV-No n=35; CMV-Yes n=21; CMV+No n=84; CMV+Yes n=44. Each data point indicates an individual participant. Data are presented as box and whisker plots, minimum to maximum, with the center line at the median. Associations between CMV serostatus and prior COVID-19 were assessed by two-way ANOVA, with Tukey’s test post-hoc analysis of significant main effects and interactions. *\*p<0*.*05, **p<0*.*01, ***p<0*.*001, ****p<0*.*0001*.

In summary, there are significant changes to the relative composition and phenotype of peripheral blood CD8^+^ T cell and CD4^+^ T cell subsets between CMV seronegative and seropositive older adults, irrespective of prior COVID-19. The observed expansion of EMRA and terminally differentiated T cells, as well as reduced surface expression of the costimulatory molecule CD28 on CD8_N_ T cells in particular, may influence vaccine-specific T cell responses.

### CMV serostatus influences CD4^+^ and CD8^+^ T cell SARS-CoV-2 antigen-induced recall responses in older adults

An activation-induced marker (AIM) assay was used to examine T cell memory responses by stimulation with the SARS-CoV-2 Spike antigen after 2^nd^ and 3^rd^ dose SARS-CoV-2 mRNA vaccinations (Figure 5; Table 4). SARS-CoV-2 vaccines are unusual in that healthy adults generate strong CD4^+^ T cell memory recall responses, but weaker CD8^+^ T cell memory responses (64). We also made similar observations in older adults. Most study participants had CD4^+^ T cell responses to SARS-CoV-2 Spike (post-dose 2: 93.1%; post-dose 3: 95.9%), but only 19.5% of participants had Spike-elicited CD8^+^ memory T cell responses after 2 vaccine doses, though this increased to 29.7% of participants after 3 vaccine doses (Table 4). CMV serostatus did not influence the number of individuals with SARS-CoV-2 Spike-AIM^+^CD4^+^ T cells or Spike-AIM^+^CD8^+^ T cells after 2^nd^ or 3^rd^ dose vaccinations. Grouped analyses revealed a significant main effect of CMV serostatus on the frequency of Spike-AIM^+^CD8^+^ T cells (Figure 5A). However, despite greater variance of data in CMV seropositive individuals, post-hoc analyses by CMV serostatus were not significant post-dose 2 or post-dose 3. Intra-individual paired analyses also showed no main effects of vaccine dose nor CMV serostatus on activation of Spike-AIM^+^CD8^+^ T cells (Supplementary Figure 3A). Prior COVID-19 did not influence the prevalence of Spike-AIM^+^CD8^+^ T cells after 2 or 3 vaccine doses, though CMV seropositivity contributed to increased Spike-AIM^+^CD8^+^ T cell activation post-dose 2 (Figure 6). Grouped analyses on a population and intra-individual basis showed a main effect of vaccine dose (Figure 5B; Supplementary Figure 3B), but the prevalence of Spike-AIM^+^CD4^+^ T cells was likewise not different by CMV serostatus (Figure 5B), nor prior COVID-19 (Figure 6B&G). Therefore, CMV serostatus contributes to increased CD8^+^ T cell, but not CD4^+^ T cell, memory recall responses to the SARS-CoV-2 Spike protein several months after 2^nd^ and 3^rd^ dose vaccinations.

**Table 4.** Responders to vaccination by CMV serostatus – SARS-CoV-2 Spike T cell memory recall responses.

| T cell Population | Vaccine Doses | Total Responder Frequency |  |  | Responder Frequency by CMV Seropositivity |  |  |  |  |
| --- | --- | --- | --- | --- | --- | --- | --- | --- | --- |
|  |  | n | Frequency (%) | *p*<br>(Fisher's exact test) | Seronegative |  | Seropositive |  | *p*<br>(Fisher's exact test) |
|  |  |  |  |  | n | Frequency (%) | n | Frequency (%) |  |
| Spike-AIM<sup>+</sup>CD8<sup>+</sup> | 2 | 31/159 | 19.5 | 0.0943 | 6/50 | 12.0 | 24/108 | 22.2 | 0.1898 |
|  | 3 | 22/74 | 29.7 |  | 3/23 | 13.0 | 19/51 | 37.3 | 0.0532 |
| Spike-AIM<sup>+</sup>CD4<sup>+</sup> | 2 | 148/159 | 93.1 | 0.5570 | 45/50 | 90.0 | 102/108 | 94.4 | 0.3262 |
|  | 3 | 71/74 | 95.9 |  | 22/23 | 95.7 | 49/51 | 96.1 | >0.9999 |

**Figure 5.**
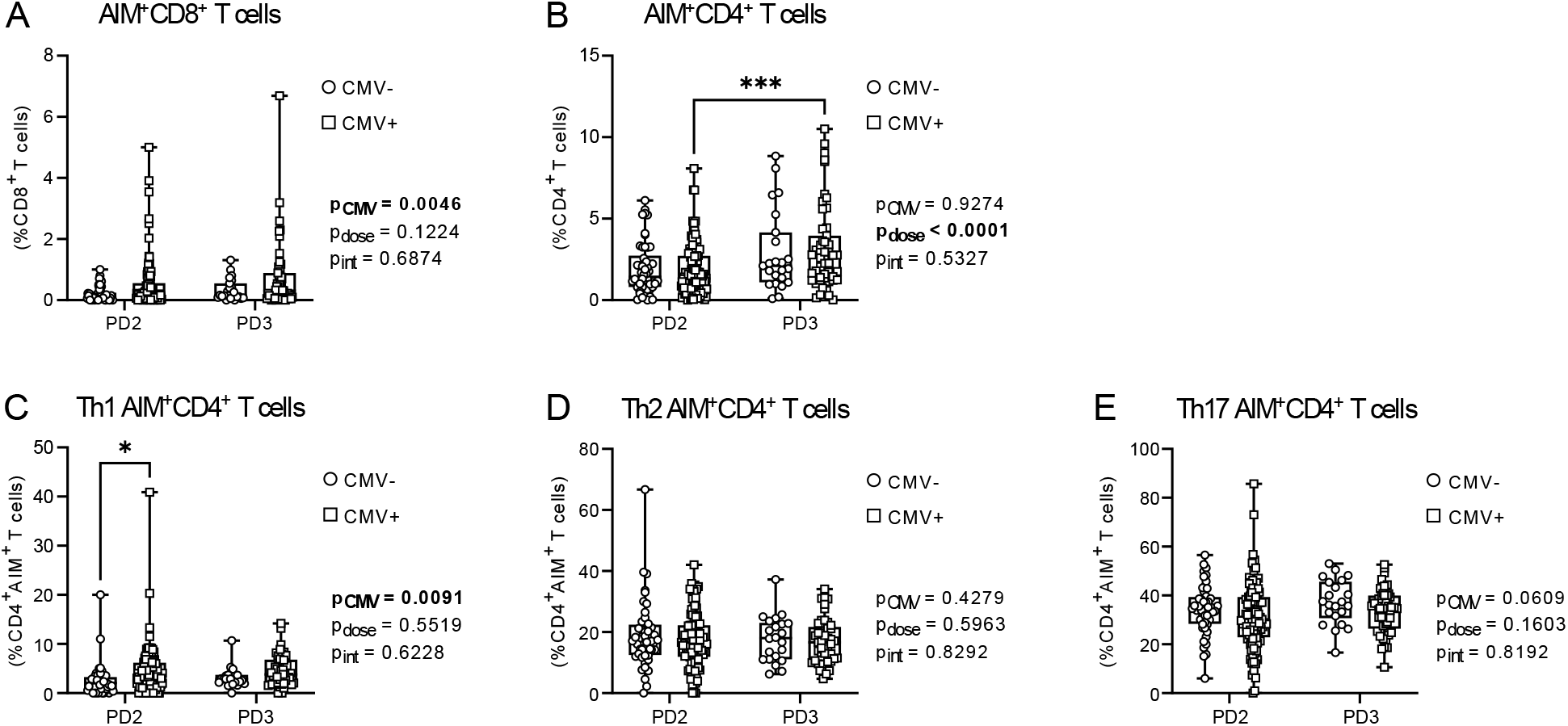
CD4^+^ and CD8^+^ T cell AIM responses to SARS-CoV-2 Spike in older adults. T cell memory responses to SARS-CoV-2 Spike were assessed by activation induced marker assay in CMV seronegative (CMV-) and CMV seropositive (CMV+) individuals post-dose 2 (PD2) and post-dose 3 (PD3) SARS-CoV-2 vaccination. (A) AIM^+^CD8^+^ T cells (expressing CD69 and CD137) were measured as a proportion of total CD8^+^ T cells. (B) AIM^+^CD4^+^ T cells (expressing CD25 and OX40) were measured as a proportion of total CD4^+^ T cells. AIM^+^CD4^+^ T cell Th1 (C), Th2 (D), and Th17 (E) subsets. Each data point indicates an individual participant. For A-B data from all individuals is graphed irrespective of whether they meet cut-off requirements for a ‘positive’ result. For C-E, only data from individuals with ‘positive’ Spike-AIM^+^CD4^+^ T cell memory recall responses were graphed (Table 4). CMV-PD2 n=51; CMV-PD3 n=23; CMV+PD2 n=108; CMV+PD3 n=51. Data are presented as box and whisker plots, minimum to maximum, with the center line at the median. Associations between CMV serostatus and vaccine dose were assessed by two-way ANOVA, with Tukey’s test post-hoc analysis of significant main effects. *\*p<0*.*05, ***p<0*.*001*.

**Figure 6.**
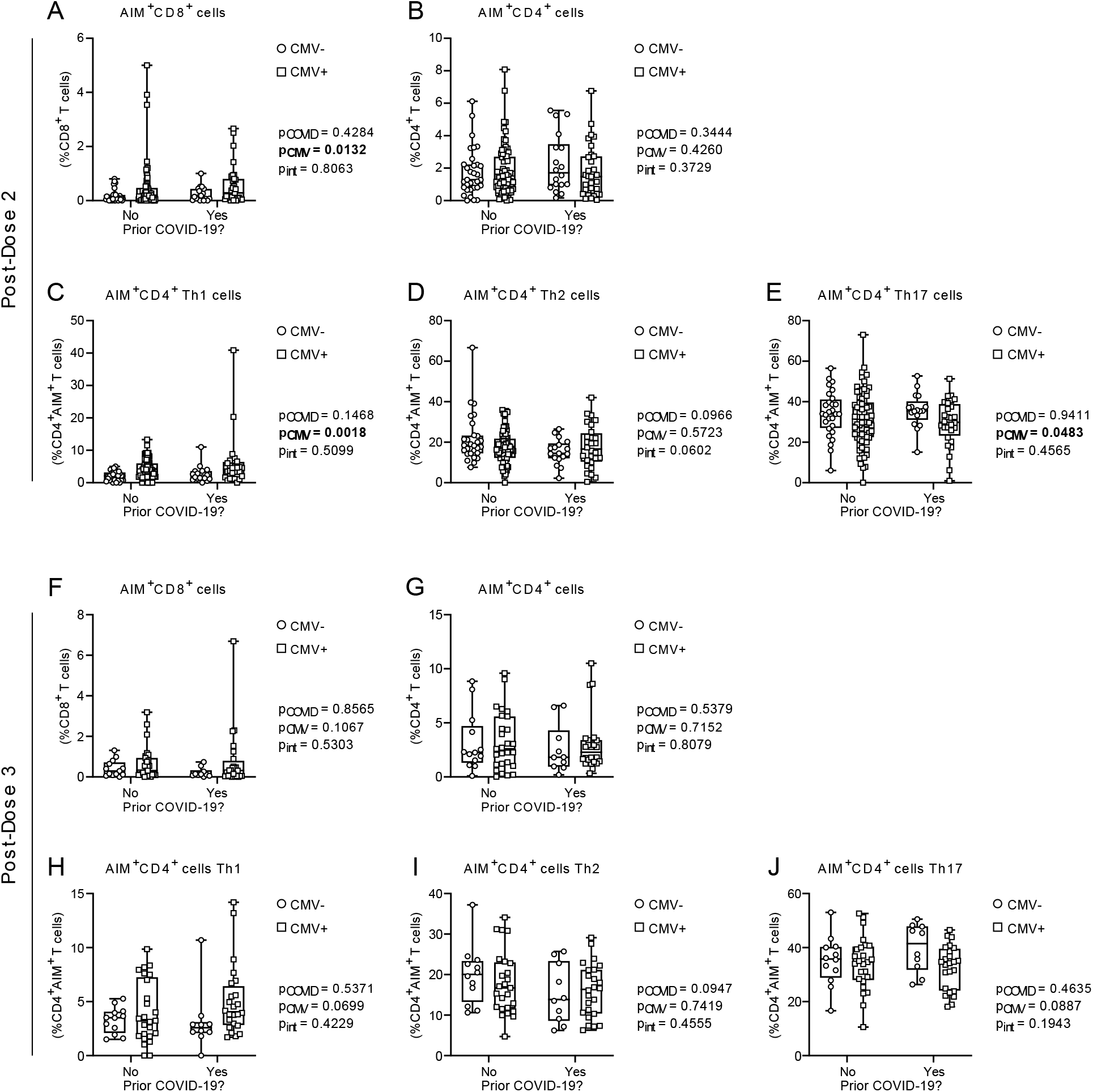
CD4^+^ and CD8^+^ T cell AIM responses to SARS-CoV-2 Spike after COVID-19 in older adults by CMV serostatus. T cell memory responses to SARS-CoV-2 Spike were assessed by activation induced marker assay in CMV seronegative (CMV-) and CMV seropositive (CMV+) individuals. Data are stratified by prior SARS-CoV-2 infection history. Post-dose 2: (A) AIM^+^CD8^+^ T cells (expressing CD69 and CD137) were measured as a proportion of total CD8^+^ T cells; (B) AIM^+^CD4^+^ T cells (expressing CD25 and OX40) were measured as a proportion of total CD4^+^ T cells; AIM^+^CD4^+^ T cell Th1 (C), Th2 (D), and Th17 (E) subsets. Post-dose 3: (F) AIM^+^CD8^+^ T cells as a proportion of total CD8^+^ T cells; (G) AIM^+^CD4^+^ T cells as a proportion of total CD4^+^ T cells; AIM^+^CD4^+^ T cell Th1 (H), Th2 (I), and Th17 (J) subsets. For C-E and H-J, only data from individuals with ‘positive’ Spike-AIM^+^CD4^+^ T cell memory recall responses were graphed (Table 4). PD2: CMV-No n=33; CMV-Yes n=18; CMV+No n=77; CMV+Yes n=31. PD3: CMV-No n=13; CMV-Yes n=10; CMV+No n=27; CMV+Yes n=24. Each data point indicates an individual participant. Data are presented as box and whisker plots, minimum to maximum, with the center line at the median. Associations between CMV serostatus and prior COVID-19 were assessed by two-way ANOVA, with Tukey’s test post-hoc analysis of significant main effects.

CD4^+^ memory T cells are comprised of a number of different functional subsets, including T helper 1 (Th1), T helper 2 (Th2), and T helper 17 (Th17) cells (65), which were further characterized. There was a significant effect of CMV serostatus on the frequency of Th1 Spike-AIM^+^CD4^+^ T cells, with post-hoc analyses showing an increase in CMV seropositive individuals after 2 but not 3 vaccine doses (Figure 5C; Supplementary Figure 3C). Th2 and Th17 Spike-AIM^+^CD4^+^ T cell frequencies were not influenced by CMV serostatus (Figure 5D-E), though paired analyses by dose and CMV serostatus suggested a significant increase in Th17 responses in CMV seropositive individuals after 3^rd^ vaccine doses (Supplementary Figure 3E). Interestingly, when we considered these data in context of prior SARS-CoV-2 infection, CMV serostatus had a main effect on the prevalence of Spike-AIM^+^CD4^+^ Th1 and Th17 T cells, which increased and decreased, respectively, with CMV seropositivity, though only post-dose 2 (Figure 6A&E), suggesting dose-dependent modulation of vaccine-associated T cell memory responses by CMV serostatus.

To determine if the observed contributions of CMV serostatus to increased AIM^+^CD8^+^ T cell and AIM^+^CD4^+^ Th1 T cell memory responses are consistent across different stimuli, we also examined T cell AIM memory responses after TCR-independent polyclonal stimulation with CytoStim, and stimulation with influenza hemagglutinin antigens (Figure 7). As we observed for SARS-CoV-2 Spike-activated memory T cells, both CytoStim and HA stimulation resulted in an increased prevalence of AIM^+^CD8^+^ T cells in CMV seropositive individuals (Figure 7A, 4F), though AIM^+^CD4^+^ T cell frequency was not affected by CMV serostatus (Figure 7B, 4G). Prior COVID-19 did not influence the frequencies of AIM^+^ T cells (Supplementary Figure 4). These data show that effects of CMV seropositivity on AIM^+^CD8^+^ and AIM^+^CD4^+^ T cell frequencies are consistent across different stimuli. CytoStim-stimulated CD4^+^ T cells, like Spike-stimulated CD4^+^ T cells, also showed distinct Th1 skewed AIM^+^CD4^+^ T cell responses, though this was not observed after HA stimulation. These data collectively suggest that CMV serostatus alters the T cell response to polyclonal activation (i.e., with CytoStim), and has differential effects on the recall response of memory T cells to specific antigens (i.e., influenza HA or SARS-CoV-2 Spike). However, CMV serostatus does not alter the ability of older adults to generate lasting CD4^+^ or CD8^+^ T cell memory, nor the incidence of T cell memory recall activation in response to SARS-CoV-2 Spike protein after mRNA vaccination.

**Figure 7.**
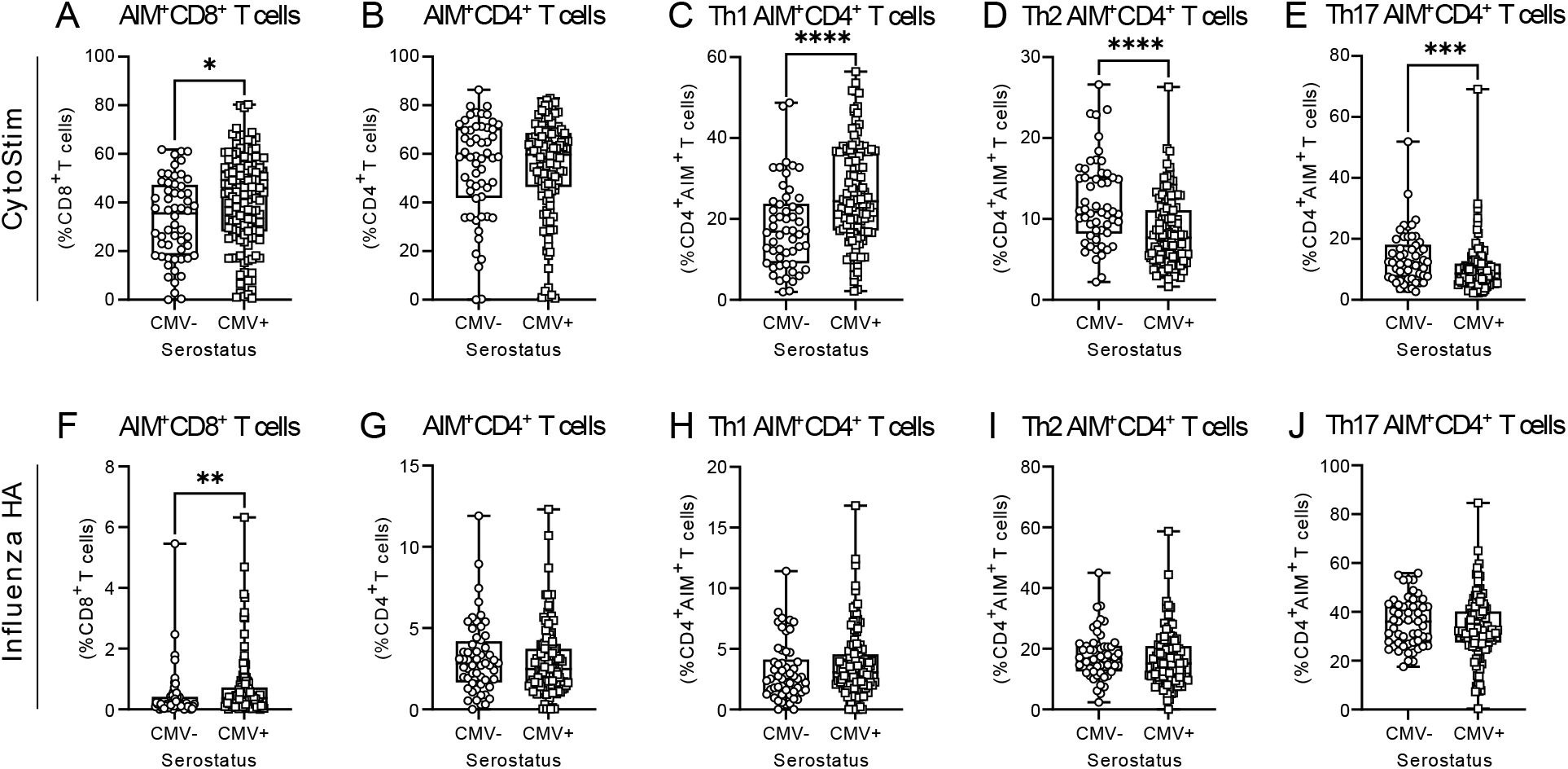
CD4^+^ and CD8^+^ T cell AIM responses to CytoStim and influenza HA in older adults. AIM assays were assessed by flow cytometry analysis after whole blood stimulation. CytoStim-induced responses: (A) AIM^+^CD8^+^ T cells (expressing CD69 and CD137) as a proportion of total CD8^+^ T cells; (B) AIM^+^CD4^+^ T cells (expressing CD25 and OX40) as a proportion of total CD4^+^ T cells; AIM^+^CD4^+^ T cell Th1 (C), Th2 (D), and Th17 (E) subsets. Influenza HA-induced responses: (F) AIM^+^CD8^+^ T cells as a proportion of total CD8^+^ T cells; (G) AIM^+^CD4^+^ T cells as a proportion of total CD4^+^ T cells; AIM^+^CD4^+^ T cell Th1 (H), Th2 (I), and Th17 (J) subsets. Each data point indicates an individual participant. Data is pooled from all blood collections, and for participants with two different collection time points, data are only included from post-dose 2 assessments. CMV-n=57; CMV+ n=130. For C-E, only data from individuals with >5% CytoStim AIM^+^CD4^+^ T cells were graphed. For H-J, only data from individuals with >0% influenza HA AIM^+^CD4^+^ T cells were graphed. Data are presented as box and whisker plots, minimum to maximum, with the center line at the median. Associations between T cell responses and CMV serostatus were assessed by Student’s t test with Welch’s correction or Mann-Whitney U test, according to normality. *\*p<0*.*05, **p<0*.*01, ***p<0*.*001, ****p<0*.*0001*.

## Discussion

Our data suggest that despite being a significant modifier of peripheral blood T cell composition and phenotype, CMV seropositivity in older adults does not have a negative impact on the longevity of vaccine-elicited antibody quantity and quality, or T cell memory recall responses, several months after second or third doses of SARS-CoV-2 mRNA vaccines. Yet, we found that there were subtle changes in antibody and cellular responses in CMV seropositive individuals between vaccine doses and in individuals with prior COVID-19. Our study cohort included participants from multiple assisted living facilities, and it did not exclude individuals with particular health conditions (e.g., cancer, diabetes, cardiovascular disease, autoimmune disorders) or prescribed medications (e.g., immune modulating drugs). Thus, any observed effects of CMV needed to be sufficiently robust to overcome potential effects of those other factors.

Our observations of changes in the peripheral T cell repertoire in CMV seropositive individuals, and CD4^+^ and CD8^+^ T cell expression of CD28 and CD57, are consistent with prior publications that reported expansion of exhausted CD4_EMRA_ and CD8_EMRA_ T cells in CMV seropositive healthy community-dwelling adults (41, 43). There are conflicting reports as to whether CMV seropositivity is associated with a reduction in naïve CD4^+^ and CD8^+^ T cells. In this investigation, we observed similar numbers of CD4_N_ and CD8_N_ T cells in seropositive and seronegative individuals, and in particular similar expression of CD57 and CD28 on CD4_N_ T cells. Our data therefore suggest that CMV seropositivity does not influence the availability nor capacity of circulating naïve T cells to respond and generate memory responses to antigens from novel viruses like SARS-CoV-2 in older adults.

CMV seropositivity in older adults has been associated with lower frequencies of memory T cells in response to seasonal influenza, though acute infection T cell responses were unchanged (66), and there is conflicting data as to whether CMV seropositivity enhances or impairs influenza virus-specific T cell responses (33, 41, 67, 68). We found that T cell memory recall responses to influenza HA, and moreover SARS-CoV-2 Spike, were similar in CMV seropositive and seronegative individuals. Our findings are concordant with observations from a previous study that found CMV serostatus did not alter the ability of older adults to generate memory responses to the (at the time) newly emergent West Nile virus (48). Furthermore, it has been reported that T cell memory responses to the SARS-CoV-2 Spike protein are boosted in convalescent younger adults after vaccination (69, 70). We identified an increase in Spike-specific CD4^+^ T cell memory responses between 2 and 3 mRNA vaccine doses in our older adult cohort, but we did not observe increased CD8^+^ or CD4^+^ T cell memory responses in convalescent older adults after vaccination. This may in part be because our analyses of effects of prior COVID-19 were not restricted to a particular time frame post-SARS-CoV-2 infection. It is important to acknowledge that there may be early effects of CMV seropositivity on the initial generation of antibodies or cellular memory that are not apparent at the assessed time points in this study, which were several months since vaccination. Irrespective, these findings suggest that while combined effects of infection and subsequent vaccination may differ by age, older adults can elicit memory T cell responses to infection and vaccination against newly emergent viruses like SARS-CoV-2.

We observed a distinct Th1 bias after polyclonal T cell activation, and in response to the immunodominant regions of the Spike antigen after 2 doses, but not 3 doses, of SARS-CoV-2 mRNA vaccines. Th1 skewing of the cellular immune response after influenza virus vaccination has been previously noted in CMV seropositive infants and young adults and mice (68, 71), as well as in older adults (72). Strong Th1 CD4^+^ T cell responses have been associated with lower disease severity in unvaccinated COVID-19 patients (73), and SARS-CoV-2 Spike-elicited CD4^+^ T cell memory responses in unvaccinated convalescent individuals have also been identified to be primarily Th1-differentiated (69, 70). However, we did not identify a main effect of prior COVID-19, nor an interaction of CMV serostatus and prior COVID-19, on the prevalence of CD4^+^ T cell Th1 responses in vaccinated older adults. These data indicate that in older adults CMV seropositivity is associated with Th1-biased CD4^+^ T cell responses, which are not further modified by prior SARS-CoV-2 infection. Our observations also suggest that Spike-specific T cell memory recall responses change between 2 and 3 vaccine doses in older adults, congruent with observations of changes in memory T cell phenotype between vaccine doses in younger adults (69).

Our data also show that CMV seropositivity does not prevent production of anti-Spike or anti-RBD IgG, IgA, or IgM antibodies after SARS-CoV-2 mRNA, though we did observe interaction effects between CMV seropositivity and prior COVID-19 both post-dose 2 and post-dose 3 for anti-Spike and anti-RBD IgA antibodies, and post-dose 2 for IgM antibodies. These interactions may be reflective of our limited sample size, or differences in time since infection between CMV seropositive and CMV seronegative individuals. CMV seropositive individuals have been reported to have increased B cell proliferation and mutations within the immunoglobulin heavy chain sequences of IgM and IgG, but not IgA, isotypes (74). CMV serostatus could also have a larger effect on maturation of the antibody response via isotype switching, and thus isotype composition after viral infection, which may contribute to our observations, but to our knowledge this has not been extensively explored. Furthermore, our observations are from individuals vaccinated with mRNA vaccines, which were predominately used in older adults in assisted living facilities in Ontario, Canada. Effects of CMV-associated immune dysfunction on the quality and durability of humoral and cellular immune responses after vaccination may be ameliorated with mRNA vaccines, but other vaccine platforms (e.g., inactivated or attenuated whole virus, viral vector, or protein subunit) may have differential outcomes, which should be investigated in future studies.

Early in the pandemic, CMV seropositivity was associated with increased risk of hospitalization in COVID-19 patients (75), and CMV reactivation was later reported to have a higher incidence in patients in intensive care (76). More recently it was reported that unvaccinated individuals with latent CMV, irrespective of anti-CMV antibody levels, age, and sex, are at higher risk of SARS-CoV-2 infection and hospitalization (77). In particular, the exhausted T cells present in CMV seropositive individuals have been predicted to contribute to more severe COVID-19 pathophysiology (78). Our data does not preclude the possibility that CMV-associated remodelling of innate and adaptive immunity in older adults may contribute to the pathogenesis and severity of SARS-CoV-2 infection. CMV serostatus may in addition impact humoral or cellular protection after vaccination against breakthrough infections with current or emerging variants of concern.

In conclusion, our data shows that CMV serostatus alters the T cell repertoire but does not blunt durability of vaccine-elicited cellular memory responses nor humoral responses after 2 and 3 doses of SARS-CoV-2 mRNA vaccines in older adults in assisted living facilities. Further research is necessary to disentangle the more subtle effects of CMV serostatus on immunity after vaccination, as well as to assess its role in risk of breakthrough SARS-CoV-2 infections.

## Data Availability

Data are not available without prior approval from the Hamilton Integrated Research Ethics Board (HiREB).

## Acknowledgements

Data in this study were collected by the COVID-in-LTC Study Group. Other members of the COVID-in-LTC Study Group include Eric D. Brown, Kevin Brown, David C. Bulir, George A. Heckman, Michael P. Hillmer, John P. Hirdes, Aaron Jones, Mark Loeb, Janet E. McElhaney (posthumous), Nathan M. Stall, Parminder Raina, Marek Smieja, Ahmad Von Schlegel, Kevin Stinson, Arthur Sweetman, Chris Verschoor, and Gerry Wright. We acknowledge administrative and technical assistance from Tara Kajaks, Ahmad Rahim, Megan Hagerman, Braeden Cowbrough, Lucas Bilaver, Komal Aryal, Leslie Tan, Sussan Kianpour, Jodie Turner, and Sheneice Joseph, of the COVID-in-LTC Study Group at McMaster University. We would like to thank our participants, their families, as well as staff, for their support of this study.

## References

1. Kline, K. A., and D. M. Bowdish. 2016. Infection in an aging population. Curr. Opin. Microbiol. 29: 63–67.

2. Lord JM. 2013. The effect of ageing of the immune system on vaccination responses. Hum. Vaccin. Immunother. 9(6):1364–1367.

3. Banerjee, A., L. Pasea, S. Harris, A. Gonzalez-Izquierdo, A. Torralbo, L. Shallcross, M. Noursadeghi, D. Pillay, N. Sebire, C. Holmes, C. Pagel, W. K. Wong, C. Langenberg, B. Williams, S. Denaxas, and H. Hemingway. 2020. Estimating excess 1-year mortality associated with the COVID-19 pandemic according to underlying conditions and age: a population-based cohort study. Lancet 395: 1715–1725.

4. Cunningham, A. L., H. Lal, M. Kovac, R. Chlibek, S.-J. Hwang, J. Díez-Domingo, O. Godeaux, M. J. Levin, J. E. McElhaney, and J. Puig-Barberà. 2016. Efficacy of the herpes zoster subunit vaccine in adults 70 years of age or older. N. Engl. J. Med. 375: 1019–1032.

5. Lal, H., A. L. Cunningham, O. Godeaux, R. Chlibek, J. Diez-Domingo, S. J. Hwang, M. J. Levin, J. E. McElhaney, A. Poder, J. Puig-Barberà, T. Vesikari, D. Watanabe, L. Weckx, T. Zahaf, and T. C. Heineman. 2015. Efficacy of an adjuvanted herpes zoster subunit vaccine in older adults. N. Engl. J. Med. 372: 2087–2096.

6. Okoli, G.N., F. Racovitan, T. Abdulwahid, C.H. Righolt, and S.M. Mahmud. 2021. Variable seasonal influenza vaccine effectiveness across geographical regions, age groups and levels of vaccine antigenic similarity with circulating virus strains: A systematic review and meta-analysis of the evidence from test-negative design studies after the 2009/10 influenza pandemic. Vaccine 39(8): 1225–1240.

7. Rondy, M., N. El Omeiri, M. G. Thompson, A. Levêque, A. Moren, and S. G. Sullivan. 2017. Effectiveness of influenza vaccines in preventing severe influenza illness among adults: A systematic review and meta-analysis of test-negative design case-control studies. J. Infect. 75: 381–394.

8. Brown, K., Stall, NM., Vanniyasingam T., et al. 2021. Early impact of Ontario’s COVID-19 vaccine rollout on long-term care home residents and health care workers. Science Briefs of the Ontario COVID-19 Science Advisory Table. 2021;2(13). 10.47326/ocsat.2021.02.13.1.0.

9. Salcher-Konrad, M., S. Smith, and A. Comas-Herrera. 2021. Emerging Evidence on Effectiveness of COVID-19 Vaccines Among Residents of Long-Term Care Facilities. J. Am. Med. Dir. Assoc. 22: 1602–1603.

10. Chung, H., S. He, S. Nasreen, M. E. Sundaram, S. A. Buchan, S. E. Wilson, B. Chen, A. Calzavara, D. B. Fell, P. C. Austin, K. Wilson, K. L. Schwartz, K. A. Brown, J. B. Gubbay, N. E. Basta, S. M. Mahmud, C. H. Righolt, L. W. Svenson, S. E. MacDonald, N. Z. Janjua, M. Tadrous, and J. C. Kwong. 2021. Effectiveness of BNT162b2 and mRNA-1273 covid-19 vaccines against symptomatic SARS-CoV-2 infection and severe covid-19 outcomes in Ontario, Canada: test negative design study. BMJ 374: 1943.

11. Breznik, J. A., A. Zhang, A. Huynh, M. S. Miller, I. Nazy, D. M. E. Bowdish, and A. P. Costa. 2021. Antibody Responses 3-5 Months Post-Vaccination with mRNA-1273 or BNT163b2 in Nursing Home Residents. J. Am. Med. Dir. Assoc. 22: 2512–2514.

12. Zhang, A., J. A. Breznik, R. Clare, I. Nazy, M. S. Miller, D. M. E. Bowdish, and A. P. Costa. 2022. Antibody Responses to Third-Dose mRNA Vaccines in Nursing Home and Assisted Living Residents. J. Am. Med. Dir. Assoc. 23: 444–446.

13. Brockman, M. A., F. Mwimanzi, H. R. Lapointe, Y. Sang, O. Agafitei, P. Cheung, S. Ennis, K. Ng, S. Basra, L. Y. Lim, F. Yaseen, L. Young, G. Umviligihozo, F. H. Omondi, R. Kalikawe, L. Burns, C. J. Brumme, V. Leung, J. S. G. Montaner, D. Holmes, M. L. DeMarco, J. Simons, R. Pantophlet, M. Niikura, M. G. Romney, and Z. L. Brumme. 2021. Reduced magnitude and durability of humoral immune responses to COVID-19 mRNA vaccines among older adults. J. Infect. Dis. 225: 1129–1140.

14. Weng, N.-p., and G. Pawelec. 2021. Validation of the effectiveness of SARS-CoV-2 vaccines in older adults in “real-world” settings. Immun. Ageing. 18: 36.

15. Fulop, T., A. Larbi, G. Dupuis, A. Le Page, E. H. Frost, A. A. Cohen, J. M. Witkowski, and C. Franceschi. 2018. Immunosenescence and Inflamm-Aging As Two Sides of the Same Coin: Friends or Foes? Front. Immunol. 8: 1960–1960.

16. Fulop, T., G. Pawelec, S. Castle, and M. Loeb. 2009. Immunosenescence and vaccination in nursing home residents. Clin. Infect. Dis. 48: 443–448.

17. Zuhair, M., G. S. A. Smit, G. Wallis, F. Jabbar, C. Smith, B. Devleesschauwer, and P. Griffiths. 2019. Estimation of the worldwide seroprevalence of cytomegalovirus: A systematic review and meta-analysis. Rev. Med. Virol. 29: e2034.

18. Pawelec, G., J. E. McElhaney, A. E. Aiello, and E. Derhovanessian. 2012. The impact of CMV infection on survival in older humans. Curr. Opin. Immunol. 24: 507–511.

19. Sylwester, A. W., B. L. Mitchell, J. B. Edgar, C. Taormina, C. Pelte, F. Ruchti, P. R. Sleath, K. H. Grabstein, N. A. Hosken, and F. Kern. 2005. Broadly targeted human cytomegalovirus-specific CD4+ and CD8+ T cells dominate the memory compartments of exposed subjects. J. Exp. Med. 202: 673–685.

20. Chidrawar, S., N. Khan, W. Wei, A. McLarnon, N. Smith, L. Nayak, and P. Moss. 2009. Cytomegalovirus-seropositivity has a profound influence on the magnitude of major lymphoid subsets within healthy individuals. Clin. Expt. Immunol. 155: 423–432.

21. Sansoni, P., R. Vescovini, F. Fagnoni, C. Biasini, F. Zanni, L. Zanlari, A. Telera, G. Lucchini, G. Passeri, D. Monti, C. Franceschi, and M. Passeri. 2008. The immune system in extreme longevity. Exp. Gerontol. 43: 61–65.

22. Goronzy, J. J., and C. M. Weyand. 2017. Successful and Maladaptive T Cell Aging. Immunity 46: 364–378.

23. Dörner, T., and A. Radbruch. 2007. Antibodies and B Cell Memory in Viral Immunity. Immunity 27: 384–392.

24. Yan, Z., H. T. Maecker, P. Brodin, U. C. Nygaard, S. C. Lyu, M. M. Davis, K. C. Nadeau, and S. Andorf. 2021. Aging and CMV discordance are associated with increased immune diversity between monozygotic twins. Immun. Ageing. 18: 5.

25. Brodin, P., V. Jojic, T. Gao, S. Bhattacharya, C. J. Angel, D. Furman, S. Shen-Orr, C. L. Dekker, G. E. Swan, A. J. Butte, H. T. Maecker, and M. M. Davis. 2015. Variation in the human immune system is largely driven by non-heritable influences. Cell 160: 37–47.

26. Mekker, A., V. S. Tchang, L. Haeberli, A. Oxenius, A. Trkola, and U. Karrer. 2012. Immune senescence: relative contributions of age and cytomegalovirus infection. PLoS Pathog. 8: e1002850.

27. Olsson, J., A. Wikby, B. Johansson, S. Löfgren, B. O. Nilsson, and F. G. Ferguson. 2000. Age-related change in peripheral blood T-lymphocyte subpopulations and cytomegalovirus infection in the very old: the Swedish longitudinal OCTO immune study. Mech. Ageing Dev. 121: 187–201.

28. Crooke, S. N., I. G. Ovsyannikova, G. A. Poland, and R. B. Kennedy. 2019. Immunosenescence and human vaccine immune responses. Immun. Ageing. 16: 25.

29. Kadambari, S., P. Klenerman, and A. J. Pollard. 2020. Why the elderly appear to be more severely affected by COVID-19: The potential role of immunosenescence and CMV. Rev. Med. Virol. 30: e2144.

30. Moss, P. 2020. “The ancient and the new”: is there an interaction between cytomegalovirus and SARS-CoV-2 infection? Immun. Ageing. 17: 14.

31. Söderberg-Nauclér, C. 2021. Does reactivation of cytomegalovirus contribute to severe COVID-19 disease? Immun. Ageing. 18: 12.

32. Chen, Y., S. L. Klein, B. T. Garibaldi, H. Li, C. Wu, N. M. Osevala, T. Li, J. B. Margolick, G. Pawelec, and S. X. Leng. 2021. Aging in COVID-19: Vulnerability, immunity and intervention. Ageing Res. Rev. 65: 101205.

33. Merani, S., G. Pawelec, G. A. Kuchel, and J. E. McElhaney. 2017. Impact of Aging and Cytomegalovirus on Immunological Response to Influenza Vaccination and Infection. Front. Immunol. 8: 784.

34. Cicin-Sain, L., J. D. Brien, J. L. Uhrlaub, A. Drabig, T. F. Marandu, and J. Nikolich-Zugich. 2012. Cytomegalovirus infection impairs immune responses and accentuates T-cell pool changes observed in mice with aging. PLoS Pathog. 8: e1002849.

35. Smithey, M. J., G. Li, V. Venturi, M. P. Davenport, and J. Nikolich-Žugich. 2012. Lifelong persistent viral infection alters the naive T cell pool, impairing CD8 T cell immunity in late life. J. Immunol. 189: 5356–5366.

36. Khan, N., A. Hislop, N. Gudgeon, M. Cobbold, R. Khanna, L. Nayak, A. B. Rickinson, and P. A. Moss. 2004. Herpesvirus-specific CD8 T cell immunity in old age: cytomegalovirus impairs the response to a coresident EBV infection. J. Immunol. 173: 7481–7489.

37. Barton, E. S., D. W. White, J. S. Cathelyn, K. A. Brett-McClellan, M. Engle, M. S. Diamond, V. L. Miller, and H. W. t. Virgin. 2007. Herpesvirus latency confers symbiotic protection from bacterial infection. Nature 447: 326–329.

38. Terrazzini, N., M. Bajwa, S. Vita, D. Thomas, H. Smith, R. Vescovini, P. Sansoni, and F. Kern. 2014. Cytomegalovirus infection modulates the phenotype and functional profile of the T-cell immune response to mycobacterial antigens in older life. Exp. Gerontol. 54: 94–100.

39. Pera, A., C. Campos, A. Corona, B. Sanchez-Correa, R. Tarazona, A. Larbi, and R. Solana. 2014. CMV latent infection improves CD8+ T response to SEB due to expansion of polyfunctional CD57+ cells in young individuals. PloS One 9: e88538.

40. Smithey, M. J., V. Venturi, M. P. Davenport, A. S. Buntzman, B. G. Vincent, J. A. Frelinger, and J. Nikolich-Žugich. 2018. Lifelong CMV infection improves immune defense in old mice by broadening the mobilized TCR repertoire against third-party infection. Proc. Natl. Acad. Sci. USA 115: E6817–E6825.

41. Derhovanessian, E., H. Theeten, K. Hähnel, P. Van Damme, N. Cools, and G. Pawelec. 2013. Cytomegalovirus-associated accumulation of late-differentiated CD4 T-cells correlates with poor humoral response to influenza vaccination. Vaccine 31: 685–690.

42. Saurwein-Teissl, M., T. L. Lung, F. Marx, C. Gschösser, E. Asch, I. Blasko, W. Parson, G. Böck, D. Schönitzer, and E. Trannoy. 2002. Lack of antibody production following immunization in old age: association with CD8+ CD28– T cell clonal expansions and an imbalance in the production of Th1 and Th2 cytokines. J. Immunol. 168: 5893–5899.

43. Frasca, D., A. Diaz, M. Romero, A. M. Landin, and B. B. Blomberg. 2015. Cytomegalovirus (CMV) seropositivity decreases B cell responses to the influenza vaccine. Vaccine 33: 1433–1439.

44. Trzonkowski, P., J. Myśliwska, E. Szmit, J. Wieckiewicz, K. Lukaszuk, L. B. Brydak, M. Machała, and A. Myśliwski. 2003. Association between cytomegalovirus infection, enhanced proinflammatory response and low level of anti-hemagglutinins during the anti-influenza vaccination--an impact of immunosenescence. Vaccine 21: 3826–3836.

45. Bowyer, G., H. Sharpe, N. Venkatraman, P. B. Ndiaye, D. Wade, N. Brenner, A. Mentzer, C. Mair, T. Waterboer, T. Lambe, T. Dieye, S. Mboup, A. V. S. Hill, and K. J. Ewer. 2020. Reduced Ebola vaccine responses in CMV+ young adults is associated with expansion of CD57+KLRG1+ T cells. J. Exp. Med. 217: e20200004.

46. van den Berg, S. P. H., K. Warmink, J. A. M. Borghans, M. J. Knol, and D. van Baarle. 2019. Effect of latent cytomegalovirus infection on the antibody response to influenza vaccination: a systematic review and meta-analysis. Med. Microbiol. Immunol. 208: 305–321.

47. Sharpe, H. R., N. M. Provine, G. S. Bowyer, P. Moreira Folegatti, S. Belij-Rammerstorfer, A. Flaxman, R. Makinson, A. V. S. Hill, K. J. Ewer, A. J. Pollard, P. Klenerman, S. Gilbert, and T. Lambe. 2022. CMV-associated T cell and NK cell terminal differentiation does not affect immunogenicity of ChAdOx1 vaccination. JCI Insight 7: e154187.

48. Verschoor, C. P., V. Kohli, and C. Balion. 2018. A comprehensive assessment of immunophenotyping performed in cryopreserved peripheral whole blood. Cytometry B Clin. Cytom. 94: 662–670.

49. Ontario Ministry of Health. 2021. COVID-19 vaccine third dose recommendations. Toronto, ON.

50. Kennedy, A. E., L. Cook, J. A. Breznik, B. Cowbrough, J. G. Wallace, A. Huynh, J. W. Smith, K. Son, H. Stacey, J. Ang, A. McGeer, B. L. Coleman, M. Larché, M. Larché, N. Hambly, P. Nair, K. Ask, M. S. Miller, J. Bramson, M. K. Levings, I. Nazy, S. Svenningsen, M. Mukherjee, and D. M. E. Bowdish. 2021. Lasting Changes to Circulating Leukocytes in People with Mild SARS-CoV-2 Infections. Viruses 13: 2239.

51. Loukov, D., S. Karampatos, M. R. Maly, and D. M. E. Bowdish. 2018. Monocyte activation is elevated in women with knee-osteoarthritis and associated with inflammation, BMI and pain. Osteoarthr. Cartil. 26: 255–263.

52. Larbi, A., and T. Fulop. 2014. From “truly naïve” to “exhausted senescent” T cells: when markers predict functionality. Cytom., : j. Int. Soc. Anal. Cytol. 85: 25–35.

53. Zaunders, J. J., M. L. Munier, N. Seddiki, S. Pett, S. Ip, M. Bailey, Y. Xu, K. Brown, W. B. Dyer, M. Kim, R. de Rose, S. J. Kent, L. Jiang, S. N. Breit, S. Emery, A. L. Cunningham, D. A. Cooper, and A. D. Kelleher. 2009. High levels of human antigen-specific CD4+ T cells in peripheral blood revealed by stimulated coexpression of CD25 and CD134 (OX40). J. Immunol. 183: 2827–2836.

54. Seddiki, N., L. Cook, D. C. Hsu, C. Phetsouphanh, K. Brown, Y. Xu, S. J. Kerr, D. A. Cooper, C. M. Munier, S. Pett, J. Ananworanich, J. Zaunders, and A. D. Kelleher. 2014. Human antigen-specific CD4(+) CD25(+) CD134(+) CD39(+) T cells are enriched for regulatory T cells and comprise a substantial proportion of recall responses. Eur. J. Immunol. 44: 1644–1661.

55. Wolfl, M., J. Kuball, W. Y. Ho, H. Nguyen, T. J. Manley, M. Bleakley, and P. D. Greenberg. 2007. Activation-induced expression of CD137 permits detection, isolation, and expansion of the full repertoire of CD8+ T cells responding to antigen without requiring knowledge of epitope specificities. Blood 110: 201–210.

56. Huynh, A., D. M. Arnold, J. W. Smith, J. C. Moore, A. Zhang, Z. Chagla, B. J. Harvey, H. D. Stacey, J. C. Ang, R. Clare, N. Ivetic, V. T. Chetty, D. M. E. Bowdish, M. S. Miller, J. G. Kelton, and I. Nazy. 2021. Characteristics of Anti-SARS-CoV-2 Antibodies in Recovered COVID-19 Subjects. Viruses 13: 697.

57. National Advistory Committee on Immunization. 2021. An Advisory Committee Statement (ACS) National Advisory Committee on Immunization (NACI): Guidance on booster COVID-19 vaccine doses in Canada - Update December 3, 2021. Public Health Agency of Canada.

58. Hirabara, S. M., T. D. A. Serdan, R. Gorjao, L. N. Masi, T. C. Pithon-Curi, D. T. Covas, R. Curi, and E. L. Durigon. 2021. SARS-COV-2 Variants: Differences and Potential of Immune Evasion. Front. Cell. Infect. Microbiol. 11: 781429.

59. Klenerman, P., and A. Oxenius. 2016. T cell responses to cytomegalovirus. Nat. Rev. Immunol. 16: 367–377.

60. Strioga, M., V. Pasukoniene, and D. Characiejus. 2011. CD8+ CD28– and CD8+ CD57+ T cells and their role in health and disease. Immunol. 134: 17–32.

61. Fletcher, J. M., M. Vukmanovic-Stejic, P. J. Dunne, K. E. Birch, J. E. Cook, S. E. Jackson, M. Salmon, M. H. Rustin, and A. N. Akbar. 2005. Cytomegalovirus-specific CD4+ T cells in healthy carriers are continuously driven to replicative exhaustion. J. Immunol. 175: 8218–8225.

62. Henson, S. M., N. E. Riddell, and A. N. Akbar. 2012. Properties of end-stage human T cells defined by CD45RA re-expression. Curr. Opin. Immunol. 24: 476–481.

63. Brenchley, J. M., N. J. Karandikar, M. R. Betts, D. R. Ambrozak, B. J. Hill, L. E. Crotty, J. P. Casazza, J. Kuruppu, S. A. Migueles, and M. Connors. 2003. Expression of CD57 defines replicative senescence and antigen-induced apoptotic death of CD8+ T cells. Blood 101: 2711–2720.

64. Neidleman, J., X. Luo, M. McGregor, G. Xie, V. Murray, W. C. Greene, S. A. Lee, and N. R. Roan. 2021. mRNA vaccine-induced T cells respond identically to SARS-CoV-2 variants of concern but differ in longevity and homing properties depending on prior infection status. eLife 10: e72619.

65. Crotty, S. 2015. A brief history of T cell help to B cells. Nat. Rev. Immunol. 15: 185–189.

66. van den Berg, S. P. H., J. Lanfermeijer, R. H. J. Jacobi, M. Hendriks, M. Vos, R. van Schuijlenburg, N. M. Nanlohy, J. A. M. Borghans, J. van Beek, D. van Baarle, and J. de Wit. 2021. Latent CMV Infection Is Associated With Lower Influenza Virus-Specific Memory T-Cell Frequencies, but Not With an Impaired T-Cell Response to Acute Influenza Virus Infection. Front. Immunol. 12: 663664.

67. Theeten, H., C. Mathei, K. Peeters, B. Ogunjimi, H. Goossens, M. Ieven, P. Van Damme, and N. Cools. 2016. Cellular Interferon Gamma and Granzyme B Responses to Cytomegalovirus-pp65 and Influenza N1 Are Positively Associated in Elderly. Viral Immunol. 29: 169–175.

68. Furman, D., V. Jojic, S. Sharma, S. S. Shen-Orr, C. J. Angel, S. Onengut-Gumuscu, B. A. Kidd, H. T. Maecker, P. Concannon, C. L. Dekker, P. G. Thomas, and M. M. Davis. 2015. Cytomegalovirus infection enhances the immune response to influenza. Sci. Transl. Med. 7: 281ra243.

69. Neidleman, J., X. Luo, J. Frouard, G. Xie, G. Gill, E. S. Stein, M. McGregor, T. Ma, A. F. George, A. Kosters, W. C. Greene, J. Vasquez, E. Ghosn, S. Lee, and N. R. Roan. 2020. SARS-CoV-2-Specific T Cells Exhibit Phenotypic Features of Helper Function, Lack of Terminal Differentiation, and High Proliferation Potential. Cell. Rep. Med. 1(6): 100081.

70. Grifoni, A., D. Weiskopf, S. I. Ramirez, J. Mateus, J. M. Dan, C. R. Moderbacher, S. A. Rawlings, A. Sutherland, L. Premkumar, R. S. Jadi, D. Marrama, A. M. de Silva, A. Frazier, A. F. Carlin, J. A. Greenbaum, B. Peters, F. Krammer, D. M. Smith, S. Crotty, and A. Sette. 2020. Targets of T Cell Responses to SARS-CoV-2 Coronavirus in Humans with COVID-19 Disease and Unexposed Individuals. Cell 181: 1489-1501.e1415.

71. Miles, D. J., M. van der Sande, D. Jeffries, S. Kaye, J. Ismaili, O. Ojuola, M. Sanneh, E. S. Touray, P. Waight, S. Rowland-Jones, H. Whittle, and A. Marchant. 2007. Cytomegalovirus infection in Gambian infants leads to profound CD8 T-cell differentiation. J. Virol. 81: 5766–5776.

72. Felismino, E. S., J. M. B. Santos, M. Rossi, C. A. F. Santos, E. L. Durigon, D. B. L. Oliveira, L. M. Thomazelli, F. R. Monteiro, A. Sperandio, J. S. Apostólico, C. N. França, J. B. Amaral, G. R. Amirato, R. P. Vieira, M. Vaisberg, and A. L. L. Bachi. 2021. Better Response to Influenza Virus Vaccination in Physically Trained Older Adults Is Associated With Reductions of Cytomegalovirus-Specific Immunoglobulins as Well as Improvements in the Inflammatory and CD8+ T-Cell Profiles. Front. Immunol. 12: 713763.

73. Rydyznski Moderbacher, C., S. I. Ramirez, J. M. Dan, A. Grifoni, K. M. Hastie, D. Weiskopf, S. Belanger, R. K. Abbott, C. Kim, J. Choi, Y. Kato, E. G. Crotty, C. Kim, S. A. Rawlings, J. Mateus, L. P. V. Tse, A. Frazier, R. Baric, B. Peters, J. Greenbaum, E. Ollmann Saphire, D. M. Smith, A. Sette, and S. Crotty. 2020. Antigen-Specific Adaptive Immunity to SARS-CoV-2 in Acute COVID-19 and Associations with Age and Disease Severity. Cell 183: 996-1012.e1019.

74. Wang, C., Y. Liu, L. T. Xu, K. J. L. Jackson, K. M. Roskin, T. D. Pham, J. Laserson, E. L. Marshall, K. Seo, J.-Y. Lee, D. Furman, D. Koller, C. L. Dekker, M. M. Davis, A. Z. Fire, and S. D. Boyd. 2014. Effects of aging, cytomegalovirus infection, and EBV infection on human B cell repertoires. J. Immunol. 192: 603–611.

75. Shrock, E., E. Fujimura, T. Kula, R. T. Timms, I. H. Lee, Y. Leng, M. L. Robinson, B. M. Sie, M. Z. Li, Y. Chen, J. Logue, A. Zuiani, D. McCulloch, F. J. N. Lelis, S. Henson, D. R. Monaco, M. Travers, S. Habibi, W. A. Clarke, P. Caturegli, O. Laeyendecker, A. Piechocka-Trocha, J. Z. Li, A. Khatri, H. Y. Chu, A. C. Villani, K. Kays, M. B. Goldberg, N. Hacohen, M. R. Filbin, X. G. Yu, B. D. Walker, D. R. Wesemann, H. B. Larman, J. A. Lederer, and S. J. Elledge. 2020. Viral epitope profiling of COVID-19 patients reveals cross-reactivity and correlates of severity. Science 370: eabd4250.

76. Simonnet, A., I. Engelmann, A. S. Moreau, B. Garcia, S. Six, A. El Kalioubie, L. Robriquet, D. Hober, and M. Jourdain. 2021. High incidence of Epstein-Barr virus, cytomegalovirus, and human-herpes virus-6 reactivations in critically ill patients with COVID-19. Infect. Dis. Now 51: 296–299.

77. Alanio, C., A. Verma, D. Mathew, S. Gouma, G. Liang, T. Dunn, D. A. Oldridge, J. E. Weaver, L. Kuri-Cervantes, M. B. Pampena, M. R. Betts, R. G. Collman, F. D. Bushman, N. J. Meyer, S. E. Hensley, D. Rader, and E. J. Wherry. 2022. Cytomegalovirus latent infection is associated with an increased risk of COVID-19-related hospitalization. J. Infect. Dis. jiac020.

78. Del Valle, D. M., S. Kim-Schulze, H.-H. Huang, N. D. Beckmann, S. Nirenberg, B. Wang, Y. Lavin, T. H. Swartz, D. Madduri, A. Stock, T. U. Marron, H. Xie, M. Patel, K. Tuballes, O. Van Oekelen, A. Rahman, P. Kovatch, J. A. Aberg, E. Schadt, S. Jagannath, M. Mazumdar, A. W. Charney, A. Firpo-Betancourt, D. R. Mendu, J. Jhang, D. Reich, K. Sigel, C. Cordon-Cardo, M. Feldmann, S. Parekh, M. Merad, and S. Gnjatic. 2020. An inflammatory cytokine signature predicts COVID-19 severity and survival. Nat. Med. 26: 1636–1643.

