## Supplemental Figures 1-4 for "CMV seropositivity in older adults changes the T cell repertoire, but does not prevent antibody or cellular responses to SARS-CoV-2 vaccination"

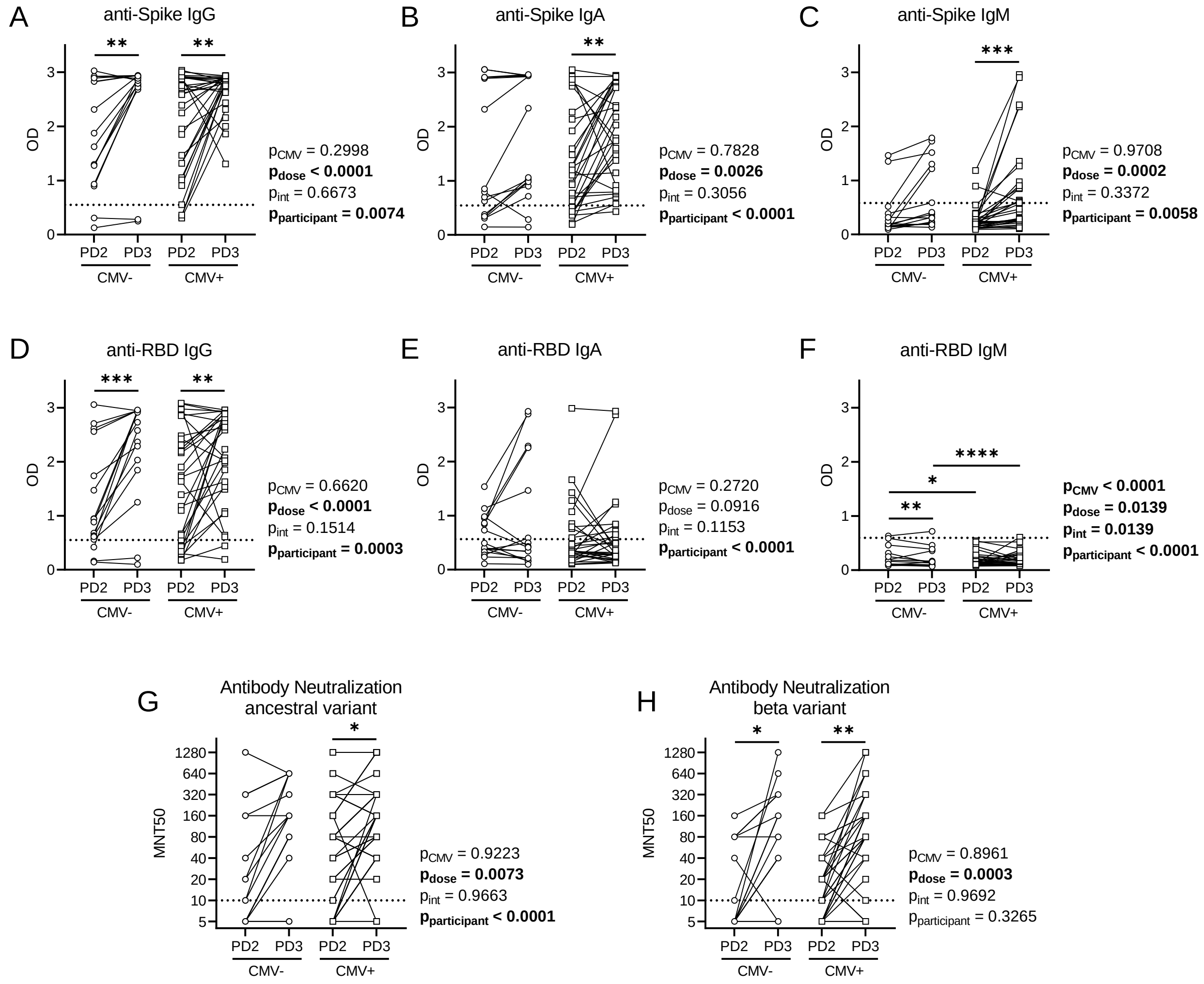


**Supplementary Figure 1. Intra-individual comparisons of antibody levels and neutralization capacity after two and three COVID-19 vaccine doses in older adults.** Serum SARS-CoV-2 anti-Spike and anti-RBD antibodies were detected by ELISA, and antibody neutralization capacity was assessed by MNT50 with live SARS-CoV-2 virus post-dose 2 (PD2) and post-dose 3 (PD3) vaccination in CMV seronegative (-ve) and CMV seropositive (+ve) individuals. Matched intra-individual data of serum antibodies: anti-Spike IgG (A), IgA (B) and IgM (C) antibodies; anti-RBD IgG (D), IgA (E), and IgM (F) antibodies. Matched intra-individual antibody neutralization capacity of: ancestral (G) and beta variant (H) SARS-CoV-2. CMV- n=17; CMV+ n=30. Dotted lines indicate the threshold of detection. Each data point indicates an individual participant and lines connect data from a single participant across post-dose 2 and post-dose 3 time points. Intra-individual associations between CMV serostatus and vaccine dose were assessed by paired two-way ANOVA, with Šidák's test post-hoc analysis of significant main effects and interactions. **p<0.05, **p<0.01, ***p<0.001, ****p<0.0001.*

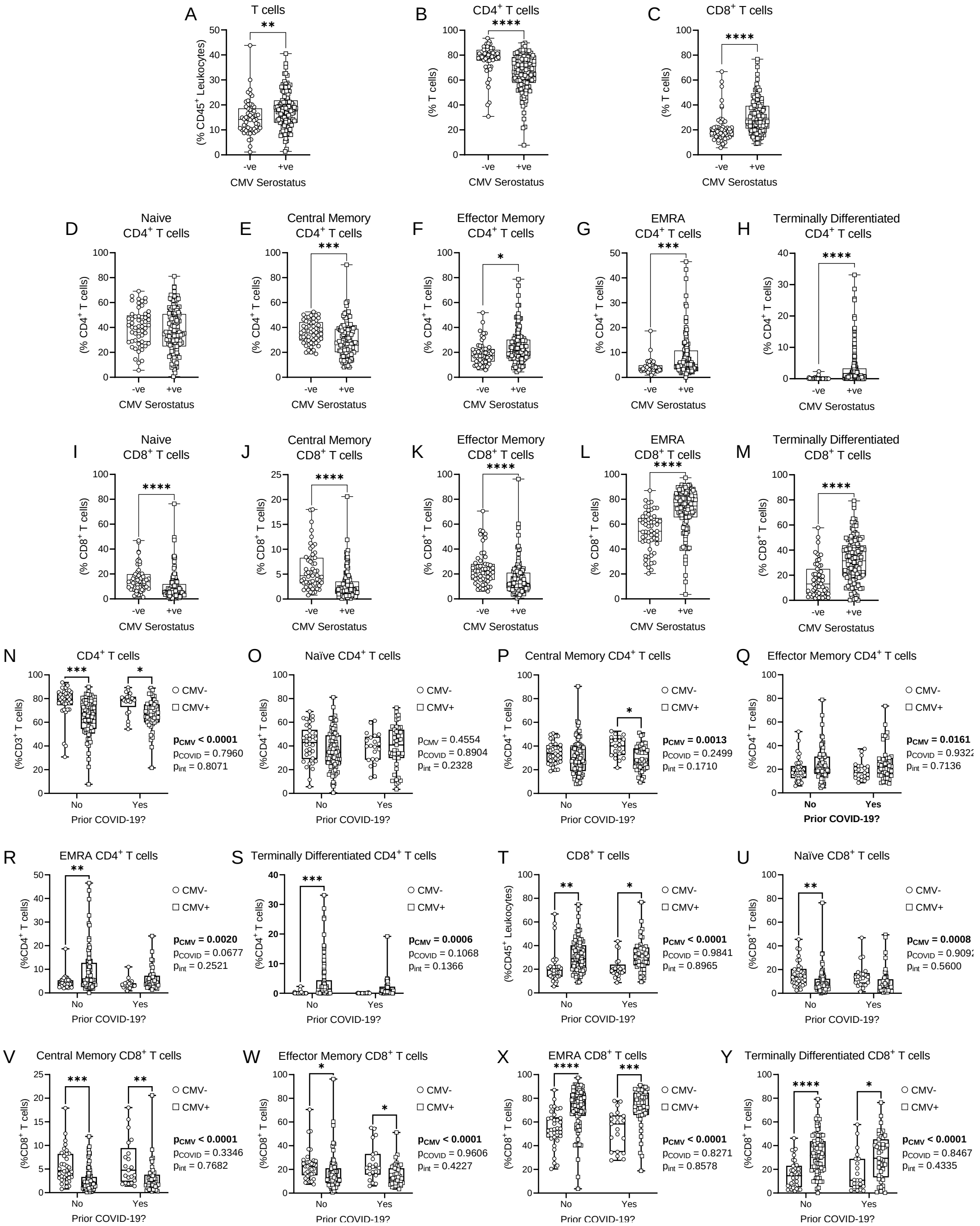


**Supplementary Figure 2. Effect of CMV serostatus and prior COVID-19 on the circulating T cell repertoire in older adults by cell frequency.**

T cell populations in whole blood were assessed by flow cytometry in CMV seronegative (-ve) and CMV seropositive (+ve) individuals, and data in N-Y are stratified by prior SARS-CoV-2 infection history (also see Figures 3 and 4). (A) Prevalence of T cells as a proportion of total leukocytes. Prevalence of: (B,N) $CD4^+$ T cells and (C,T) $CD8^+$ T cells as a proportion of total T cells. $CD4^+$ T cell subsets as a proportion of total $CD4^+$ T cells: (D,O) naïve; (E,P) central memory; (F,Q) effector memory; (G,R) EMRA; (H,S) terminally differentiated. $CD8^+$ T cell subsets as a proportion of total $CD4^+$ T cells: (I,U) naïve; (J,V) central memory; (K,W) effector memory; (L,X) EMRA; (M,Y) terminally differentiated. Blood was assessed either post-dose 2 or post-dose 3 for each participant. For A-M, CMV-ve n=56; CMV+ve n=128. For N-Y, CMV-No n=35; CMV-Yes n=21; CMV+No n=84; CMV+Yes n=44. Each data point indicates an individual participant. Data are presented as box and whisker plots, minimum to maximum, with the center line at the median. Associations between T cell subsets and CMV serostatus in A-M were assessed by Student's t test with Welch's correction or Mann-Whitney U test, according to normality. Associations between CMV serostatus and prior COVID-19 in N-Y were assessed by two-way ANOVA, with Tukey's test post-hoc analysis of significant main effects. **p<0.05, **p<0.01, ***p<0.001, ****p<0.0001.*

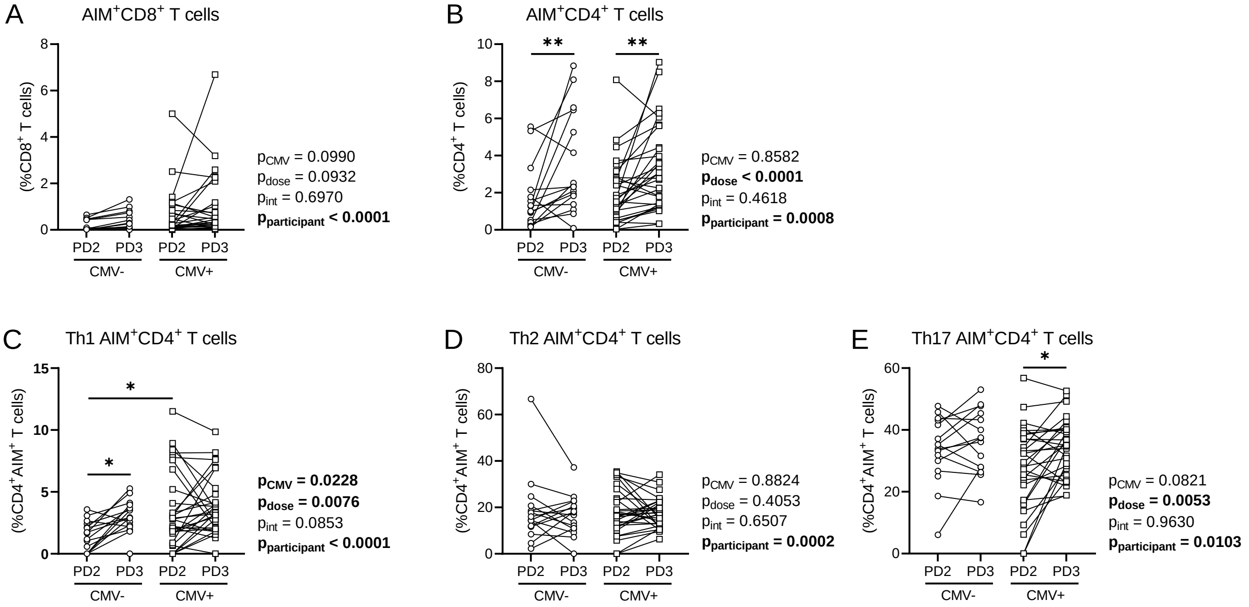


**Supplementary Figure 3. Intra-individual comparisons of CD4$^+$ and CD8$^+$ T cell AIM responses to SARS-CoV-2 Spike protein in older adults by CMV serostatus.**
T cell memory responses to SARS-CoV-2 Spike protein were assessed by activation induced marker assay in CMV seronegative (CMV-) and CMV seropositive (CMV+) individuals post-dose 2 (PD2) and post-dose 3 (PD3) SARS-CoV-2 vaccination. Matched intra-individual data are shown. (A) AIM$^+$CD8$^+$ T cells (expressing CD69 and CD137) as a proportion of total CD8$^+$ T cells. (B) AIM$^+$CD4$^+$ T cells (expressing CD25 and OX40) as a proportion of total CD4$^+$ T cells. AIM$^+$CD4$^+$ T cell Th1 (C), Th2 (D), and Th17 (E) subsets. CMV- n=17; CMV+ n=30. For C-E, only data from individuals with 'positive' Spike-AIM$^+$CD4$^+$ T cell memory recall responses were graphed (Table 4). Each data point indicates an individual participant and lines connect data from a single participant across post-dose 2 and post-dose 3 time points. Intra-individual associations between CMV serostatus and vaccine dose were assessed by paired two-way ANOVA, with Šidák's test post-hoc analysis of significant main effects. **p<0.05, **p<0.01.*

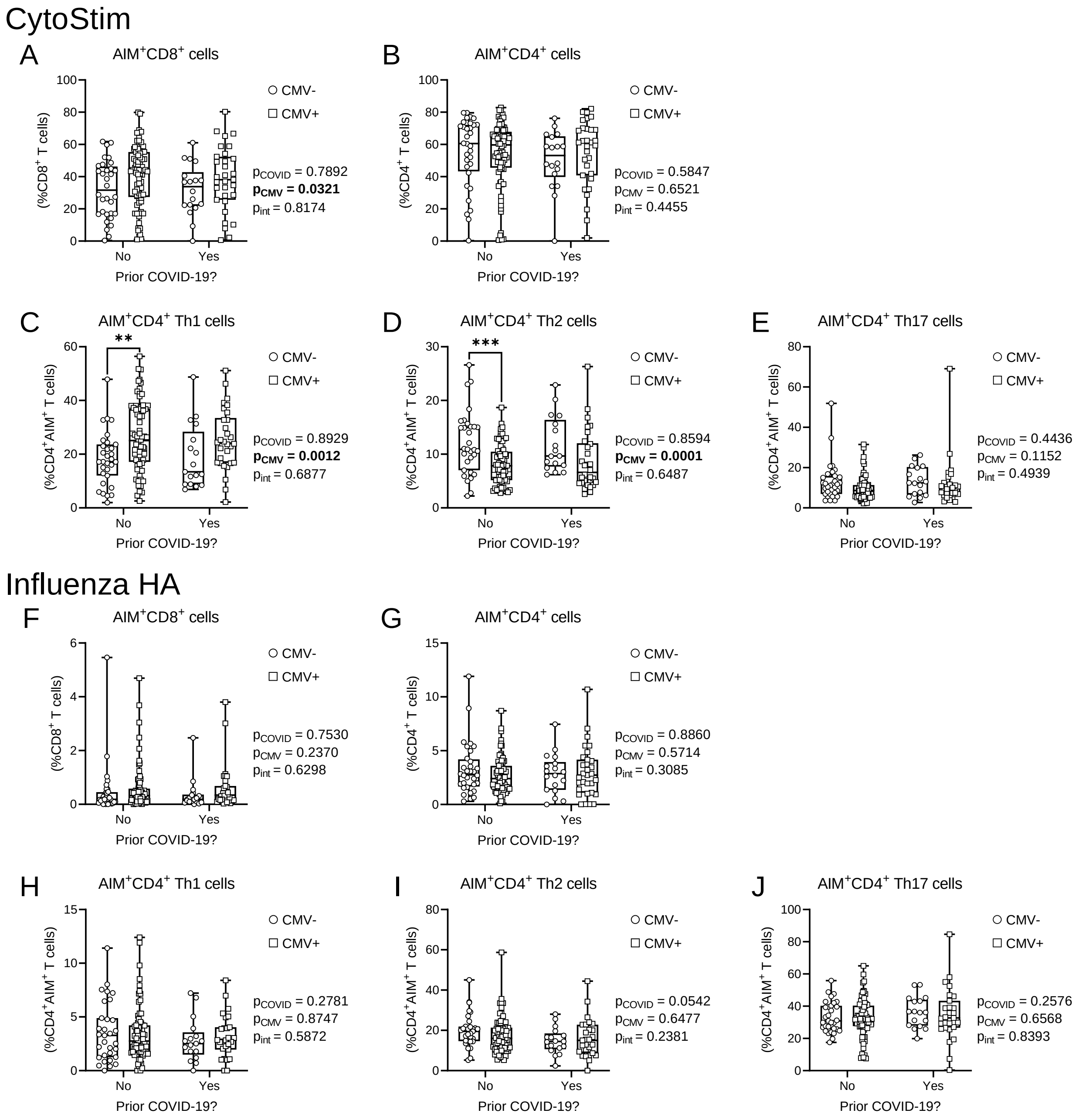


**Supplementary Figure 4. $CD4^+$ and $CD8^+$ T cell AIM responses to CytoStim and influenza HA after COVID-19 in older adults by CMV serostatus.**
T cell activation was assessed by activation induced marker assay in CMV seronegative (CMV-) and CMV seropositive (CMV+) individuals. Data are stratified by prior SARS-CoV-2 infection history. CytoStim-induced responses: (A) $AIM^+CD8^+$ T cells (expressing CD69 and CD137) as a proportion of total $CD8^+$ T cells; (B) $AIM^+CD4^+$ T cells (expressing CD25 and OX40) as a proportion of total $CD4^+$ T cells; $AIM^+CD4^+$ T cell Th1 (C), Th2 (D), and Th17 (E) subsets. Influenza HA-induced responses: (F) $AIM^+CD8^+$ T cells as a proportion of total $CD8^+$ T cells; (G) $AIM^+CD4^+$ T cells as a proportion of total $CD4^+$ T cells; $AIM^+CD4^+$ T cell Th1 (H), Th2 (I), and Th17 (J) subsets. CMV-No n=35; CMV-Yes n=21; CMV+No n=84; CMV+Yes n=44. For C-E, only data from individuals with >5% CytoStim $AIM^+CD4^+$ T cells were graphed. For H-J, only data from individuals with >0% influenza HA $AIM^+CD4^+$ T cells were graphed. Each data point indicates an individual participant. Data are presented as box and whisker plots, minimum to maximum, with the center line at the median. Associations between CMV serostatus and prior COVID-19 were assessed by two-way ANOVA, with Tukey's test post-hoc analysis of significant main effects. ***p<0.01, ***p<0.001.*
